# Genome-wide association study of anterior uveitis

**DOI:** 10.1101/2024.06.03.24308116

**Authors:** Fredrika Koskimäki, Oona Ahokas, FinnGen, Estonian Biobank Research Team, Abdelrahman Elnahas, Anu Reigo, Kadri Reis, Tõnu Esko, Priit Palta, Sanna Leinonen, Johannes Kettunen, Johanna Liinamaa, Minna K. Karjalainen, Ville Saarela

## Abstract

**Importance:** Anterior uveitis is an intraocular disease characterized by inflammation of the iris and the ciliary body and known to co-occur with autoimmune diseases. This is the largest genome-wide association study (GWAS) meta-analysis specifically for anterior uveitis to date.

**Objective:** The purpose of this study is to define genetic factors associated with anterior uveitis through genome-wide association study.

**Design, setting and participants:** In this GWAS meta-analysis we combined data from the FinnGen, Estonian Biobank and UK Biobank with a total of 12 205 anterior uveitis cases and 917 145 controls. We performed a phenome-wide association study to investigate associations across phenotypes and traits. We also evaluated genetic correlations of anterior uveitis.

**Main outcomes and measures:** Genetic variants associated with anterior uveitis.

**Results:** We identified six anterior uveitis associated loci. Genome-wide significant (*p* < 5 × 10^−8^) associations were identified for the first time at three loci (*INAVA, NLRP3* and *NOS2*). We detected associations at three loci previously reported to be associated with uveitis (*ERAP1, TNRC18* and the *HLA* region) and also replicated associations at two loci previously associated with acute anterior uveitis (*IL23R* and *HDAC2-AS2*). In phenome-wide association study, we further detected that lead single nucleotide polymorphism, SNPs, at three of the anterior uveitis associated loci (*ERAP1, INAVA* and *TNRC18*) are associated with other immunity-related phenotypes, including ankylosing spondylitis and inflammatory bowel disease. Additionally, we detected a moderate genetic correlation between anterior uveitis and inflammatory bowel disease (*r*_*g*_=0.39, *p* = 8 × 10^−5^).

**Conclusions and relevance:** We identified six anterior uveitis associated loci including three novel loci with genome-wide significance. Our findings deepen our understanding of the genetic basis of anterior uveitis and the genetic connections between anterior uveitis and immune related disorders, providing a foundation for further research and potential therapeutic interventions.

## INTRODUCTION

Anterior uveitis is an intraocular disease characterized by inflammation of the iris and the ciliary body.^1^ Non-infectious acute anterior uveitis is the most common type of uveitis presenting typically with acute unilateral pain, redness and photophobia.^1,2^ However, chronic anterior uveitis may be more insidious.^1^ Although modern immunomodulatory therapies have had a positive effect on the outcome, especially chronic anterior uveitis may lead to complications including cataract formation, macular oedema and secondary glaucoma.^3^ Anterior uveitis has been shown to co-occur with autoimmune diseases including spondyloarthropathies such as ankylosing spondylitis and psoriatic arthritis, and also with inflammatory bowel disease, juvenile idiopathic arthritis and tubulointerstitial nephritis.^4–7^

The association of anterior uveitis with human leukocyte antigen *(HLA)-B27* is well-established.^8^ Various genes have been associated with anterior uveitis in previous small-scale association studies, including immune-related genes such as those in the HLA region (*HLA-B, MICA*) and others including *CHF* and *C2, CYP27B1, KIR*, and genes from the tumor-necrosis factor family and interleukin-1 families.^9–15^ Genome-wide association studies (GWASs) have consistently detected associations at the HLA region for acute anterior uveitis.^16–18^ However, GWASs have identified only few genetic loci that may predispose to anterior uveitis, mostly due to limited sample sizes in most studies.^16–20^ Genome-wide significant associations have been found at three additional loci (*IL23R, ERAP1* and intergenic region *B3GNT2-TMEM17*) for acute anterior uveitis.^17^ Furthermore, five uveitis associated loci (*TNRC18, EID2B/SELENOV, CAVIN4, MAP6, HDAC2-AS2*) were recently reported.^18^

The purpose of this study is to find new genetic associations of anterior uveitis through genome-wide association analyses. To our knowledge, this is the largest GWAS meta-analysis specifically for anterior uveitis to date. We aim to evaluate genetic correlations of anterior uveitis and also investigate associations across phenotypes and traits through phenome-wide association analysis.

## MATERIALS AND METHODS

### Study populations

In this GWAS meta-analysis of anterior uveitis (total *n*=929 350; case *n*=12 205, control *n*=917 145), we combined data from three large population-based cohorts or biobank-related studies: The FinnGen study^20^ release 10; The Estonian Biobank (ESTBB)^21^; and The UK Biobank (UKBB)^22^. All studies comprised individuals of European origin only. Detailed study descriptions are available in Supplemental Text. In FinnGen, anterior uveitis cases were defined through hospital discharge registries (ICD10 code H20, H22* and H22.0*B96.80). In the ESTBB, cases were identified using ICD-codes (H20.0, H20.1, H20.2, H20.8, H20.9, H22.0, H22.1), while participants with other H10*-H28* codes were excluded from controls. A matching endpoint from the Pan-ancestry genetic analysis of the UK Biobank project^22^ European subsample was used.

### Genome-wide association study of anterior uveitis

GWAS was performed separately in each cohort, followed by a meta-analysis of all cohorts. In the FinnGen GWAS was conducted with a mixed-model-based whole-genome regression using the REGENIE pipeline^23^; sex, age, genotyping batches and ten principal components were included as covariates. In the ESTBB association analysis was carried out for all variants with an INFO score > 0.4 using the additive model as implemented in REGENIE v2.2.4 with standard binary trait settings.^23^ Logistic regression was carried out with adjustment for current age, age^2^, sex and 10 PCs as covariates, analyzing only variants with a minimum minor allele count of 2. In the UKBB, genome-wide genotyping was performed using the UK Biobank Axiom array and variants were imputed via the Haplotype Reference Consortium (HRC), UK10K and 1000 Genomes reference panels.^24^ To combine results of different studies in meta-analysis, an inverse variance weighted fixed-effect meta-analysis was performed. Single nucleotide polymorphisms, SNPs, with association *p*-values < 5 × 10^−8^ were considered significant, corresponding to the common threshold for genome-wide significance. Associated genomic regions were defined as follows: SNPs meeting the significance threshold were indicated as being part of the same associated locus if they were within a distance of 1 Mb. Lead SNPs were defined as the SNPs with the lowest *p*-values within each locus, and manual curation was performed to define the most likely candidate genes at each locus, using relevant resources and databases including PubMed^25^, UniProt^26^ and Gene^27^. Genomic positions refer to human genome build GRCh38.

### Comparison to previous studies

When comparing genome-wide associations in our study to previous GWASs of anterior uveitis, we defined anterior uveitis-associated loci with a distance >1 Mb from the previous associations as novel.^16– 18,20^ We considered associations with *p*-values <0.006 as evidence of replication (corresponding to Bonferroni correction with the number of previously associated loci). In addition, we performed a phenome-wide association study (PheWAS) of our lead SNPs to investigate associations across phenotypes and traits. The PheWAS was performed using PhenoScanner, v2^28^, which is a curated database of publicly available GWAS; using the NHGRI-EBI Catalog of human genome-wide association studies (GWAS catalog)^29^; and by investigating associations across FinnGen^20^ endpoints. Using PhenoScanner, we screened associations of the lead SNPs with: diseases and traits; gene expression levels; and protein levels. We screened the GWAS catalog associations with the Functional Mapping and Annotation of Genome-Wide Association Studies platform^30^ by indicating associations of SNPs in linkage disequilibrium with the lead SNPs and those of additional associated non-highly correlated SNPs (*r*^*2*^<0.6) at these loci. We screened associations with all predefined FinnGen endpoints using FinnGen Data Release R10. Genome-wide significant associations at *p*-value < 5 × 10^−8^ are reported.

### Genetic correlations

Genetic correlations were estimated using linkage disequilibrium score regression (LDSC)^31^. Publicly available GWAS summary statistics of 181 traits (diseases and anthropometric measures shown in Supplementary Table S5) obtained from the MRC IEU OpenGWAS database^32^ were used to estimate genetic correlations of anterior uveitis with these traits. We consired genetic correlations with p<0.000276 significant (corresponding to the Bonferroni correction of 181 tests).

## RESULTS

### Genome-wide association study

In this genome-wide association study, we identified six anterior uveitis associated genomic regions with genome-wide significance (Figure 1, Table 1, Supplementary Table S1)). We identified biologically plausible candidate genes in all associated loci (Table 1). These loci include three loci previously reported to be associated with anterior uveitis (*ERAP1, TNRC18* and the *HLA* region)^16–18,20^. Three novel loci were identified: *INAVA, NLRP3* and *NOS2*. SNPs at the *INAVA/KIF21B* and *NOS2* loci were previously reported to be suggestively associated with acute anterior uveitis^16,17^, and for the first time, association signal at these loci demonstrated genome-wide significant association level.

**Table 1.** Genomic regions associated with anterior uveitis.

| Lead variant<br>rsid | Genomic<br>position<br>(chr:pos) | Other<br>allele | Effect<br>allele | Nearest<br>gene | Candidate<br>gene | p-value | Odds<br>ratio | 95%<br>confidence<br>interval | Effect allele frequency<br>FinnGen/ESTBB/UKBB |
| --- | --- | --- | --- | --- | --- | --- | --- | --- | --- |
| rs6701496 | 1:201038700 | G | A | *CACNA1S* | *INAVA* | $1.03 \times 10^{-8}$ | 0.91 | 0.89-0.94 | 0.219/0.217/0.299 |
| rs145759877 | 1:248083681 | T | C | *OR2L13* | *NLRP3* | $3.69 \times 10^{-19}$ | 6.49 | 4.31-9.77 | 0.00035/0.00087/NA |
| rs572758335 | 5:96785200 | G | GA | *ERAP1* | *ERAP1* | $7.08 \times 10^{-24}$ | 0.85 | 0.83-0.88 | 0.287/NA/0.312 |
| rs116666910 | 6:31359257 | C | A | *HLA-B* | *HLA-B* | $<5 \times 10^{-324}$ | 3.29 | 3.18-3.40 | 0.080/0.064/0.043 |
| rs748670681 | 7:5397122 | C | T | *TNRC18* | *TNRC18* | $4.89 \times 10^{-40}$ | 1.61 | 1.50-1.73 | 0.0362/0.0133/NA |
| rs2301369 | 17:27802970 | G | C | *NOS2* | *NOS2* | $3.85 \times 10^{-8}$ | 1.08 | 1.05-1.11 | 0.328/0.332/0.374 |

rsID, reference SNP cluster ID; chr, chromosome; pos, position; ESTBB, Estonian BioBank; UKBB, UK BioBank; NA, not applicable, SNP not present in the data. Most significant single nucleotide polymorphism within each associated region is shown. We suggest the biologically most plausible gene within each region as the candidate gene.

**Figure 1.**
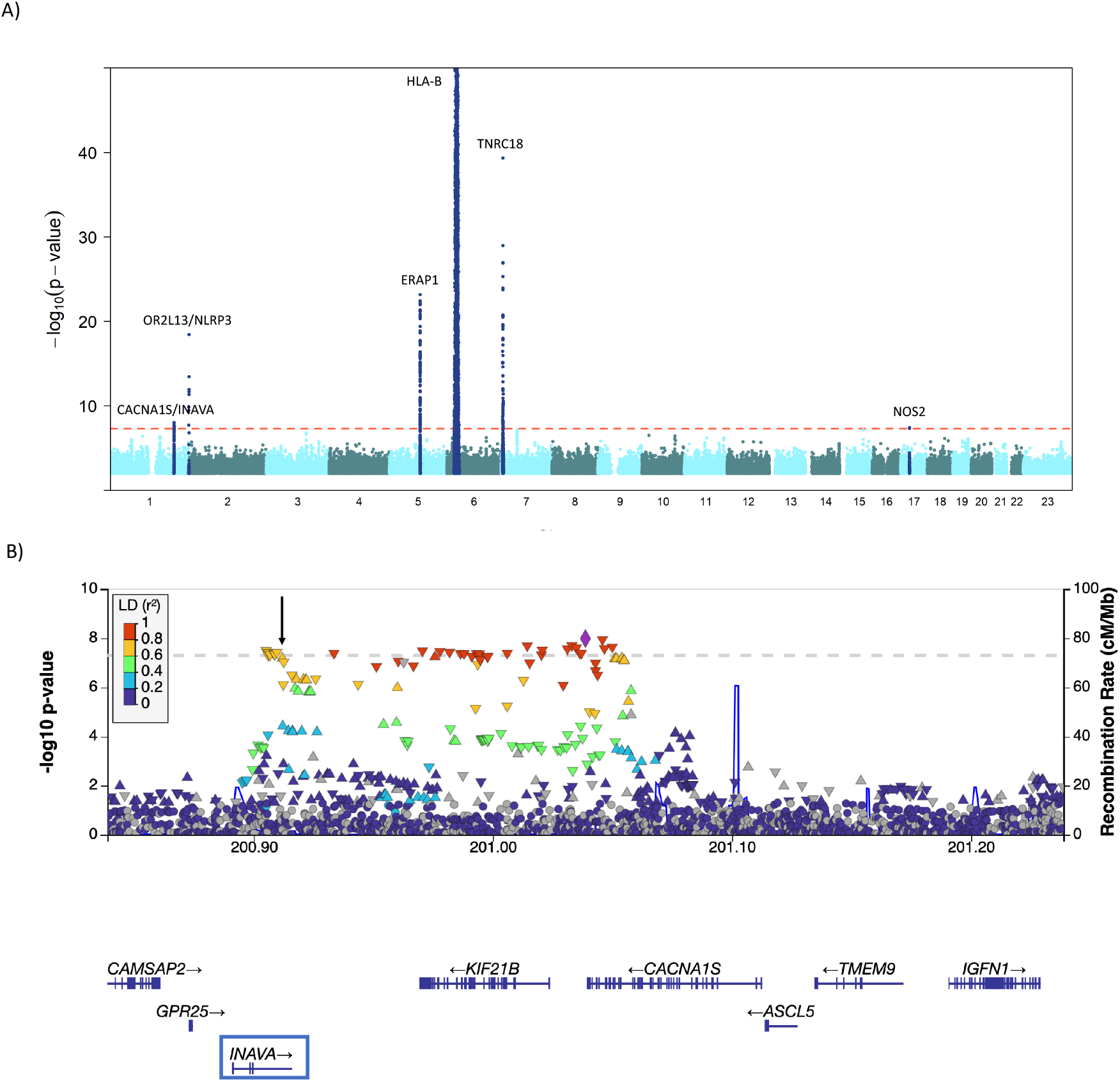
**A)** Results of genome-wide association study of anterior uveitis. GWAS results are visualized in a Manhattan plot in which each dot represents a genetic variant. X-axis shows chromosomal positions, and Y-axis represents the *p*-values on the -log10 scale. -log_10_(*p*-values) were capped at 50. The genome-wide significance threshold is marked with a dashed red line. 500 kb regions around the associated genomic regions are highlighted. **B)** Regional association plot of *INAVA* gene loci. X-axis shows genomic positions and Y-axis represents the *p*-value on the -log10 scale. European LD reference panel is applied. The genome-wide significance threshold is marked with a dashed grey line. Candidate gene is marked with a blue box. Missense SNP in linkage disequilibrium with the lead SNP is marked with a black arrow.

All novel associated loci had biologically plausible candidate genes. One of these was the *INAVA*/*KIF21B* locus in which the lead SNP rs6701496 is located near to the innate immunity activator (*INAVA*) gene (OR = 0.91, *p* = 1.03 × 10^−8^, Figure 1). This association was clearest in FinnGen (OR = 0.90, *p* = 2.99 × 10^−7^), attributed to its large size, and the effect was directionally concordant in both the Estonian Biobank (OR = 0.90) and UK Biobank (OR = 0.95) (Supplementary Figure 3). This lead SNP is in linkage disequilibrium (LD) with a missense SNP located within the *INAVA* gene (rs296520, *p* = 6.6 × 10^−8^). *INAVA* is important in innate immunity responses and it has been associated with inflammatory bowel disease.^33^ Another novel association was detected for SNP rs2301369 near the nitric oxide synthase 2 (*NOS2*) gene (OR = 1.08, *p* = 3.9 × 10^−8^). This association was similar in all three studies (FinnGen OR = 1.07, Estonian Biobank OR = 1.12, UK Biobank OR = 1.07; Supplementary Figure S3). NOS2 enzyme is involved in the production of nitric oxide in response to various inflammatory stimuli.^34^ Additionally, a novel locus was detected near *OR2L13* gene (rs145759877, OR = 6.49, *p* = 3.7 × 10^−19^). This locus primarily comprises olfactory genes. However, the signal spans a large region, and we considered the nucleotide-binding domain, leucine-rich repeat family, pyrin domain containing 3 (*NLRP3*) gene as the biologically most plausible gene (Supplementary Figure S1). This association was driven by the FinnGen data (OR = 8.44, *p* = 6.38 × 10^−19^), but also supported by the Estonian Biobank (OR = 2.83; Supplementary Figure S3). *NLRP3* gene is a part of the NLRP3 inflammasome complex and participates in the regulation of inflammation, immune responses and apoptosis.^35^

The three loci (*ERAP1, TNRC18, HLA-B*) previously associated with anterior uveitis phenotypes^16–18^ showed the most significant associations with anterior uveitis (Supplementary Figures S1 and S3). One of these associations was at the endoplasmic reticulum aminopeptidase 1 (*ERAP1*) gene (lead SNP rs572758335, *p* = 7.1 × 10^−24^). The enzyme encoded by *ERAP1* gene is important in processing peptides for presentation on MHC-I molecules, an important process for the activation and regulation of T cells.^36^ We detected an association with another previously described lead SNP, rs748670681, located in intron of the trinucleotide repeat containing 18 (*TNRC18*) gene.^20^ *TNRC18* mediates silencing of endogenous retrovirus class I and its loss activates immunity-related genes such as *IL11* and *TLR4*.^37^ The association of this SNP with several inflammatory conditions, including anterior uveitis, has been previously discussed in a study utilizing data from a previous FinnGen data freeze.^18^

### Phenome-wide association study

We conducted a phenome-wide association study of the anterior uveitis associated loci to explore the associations across phenotypes including diseases and traits, gene expression levels and protein levels (Supplementary Table S2). According to databases collecting GWAS associations (PhenoScanner and GWAS catalog), associated SNPs near the *INAVA* gene displayed associations with various immune-related diseases such as ankylosing spondylitis, celiac disease, Crohn’s disease, inflammatory bowel disease and multiple sclerosis. *INAVA* gene missense SNP (rs296520) that is in linkage disequilibrium with the lead SNP in this locus (rs6701496) was also associated with lymphocyte percentage of white cells (*p* = 2.11 × 10^−14^), inflammatory bowel disease (*p* = 3.66 × 10^−11^) and ulcerative colitis (*p* = 3.28 × 10^−8^). We detected that associated SNPs near the *ERAP1* gene showed association with multiple autoimmune diseases including acute anterior uveitis, ankylosing spondylitis and Behçet’s disease, but also with hypertension, body mass index, blood protein levels, platelet count and cardiovascular diseases. SNPs at the HLA locus were associated with hundreds of phenotypes, especially with different immune-related phenotypes. Furthermore, lead SNP at the *TNRC18* locus was associated with ulcerative colitis.

When we investigated the associations between these anterior uveitis associated loci and FinnGen endpoints (Figure 2, Supplementary Table S4), we detected that many loci were associated with inflammation and eye-related phenotypes. The lead SNP rs6701496 near *INAVA* gene was associated with inflammatory bowel disease (*p* = 1.9 × 10^−14^) and with various types of colitis, including ulcerative colitis, mucosal proctocolitis and noninfective enteritis and colitis. We also detected an association with autoimmune diseases in general. The lead SNP rs145759877 in the *OR2L13*/*NLRP3* locus was associated with eye diseases including disorders of sclera, cornea, iris and ciliary body and keratitis. We detected a significant association between the lead SNP rs748670681 near the *TNRC18* gene and inflammatory bowel disease (*p* = 2.0 × 10^−132^). Furthermore, an association was detected with specific types of inflammatory bowel diseases including ulceratice colitis, ulcerative rectosigmoiditis and Crohn’s disease of small intestine. Additionally, the same lead SNP demonstrated association with various immune-related diseases including psoriasis vulgaris and Graves disease. The lead SNP at the *HLA* region showed various highly significant associations with different immune-related conditions.

**Figure 2.**
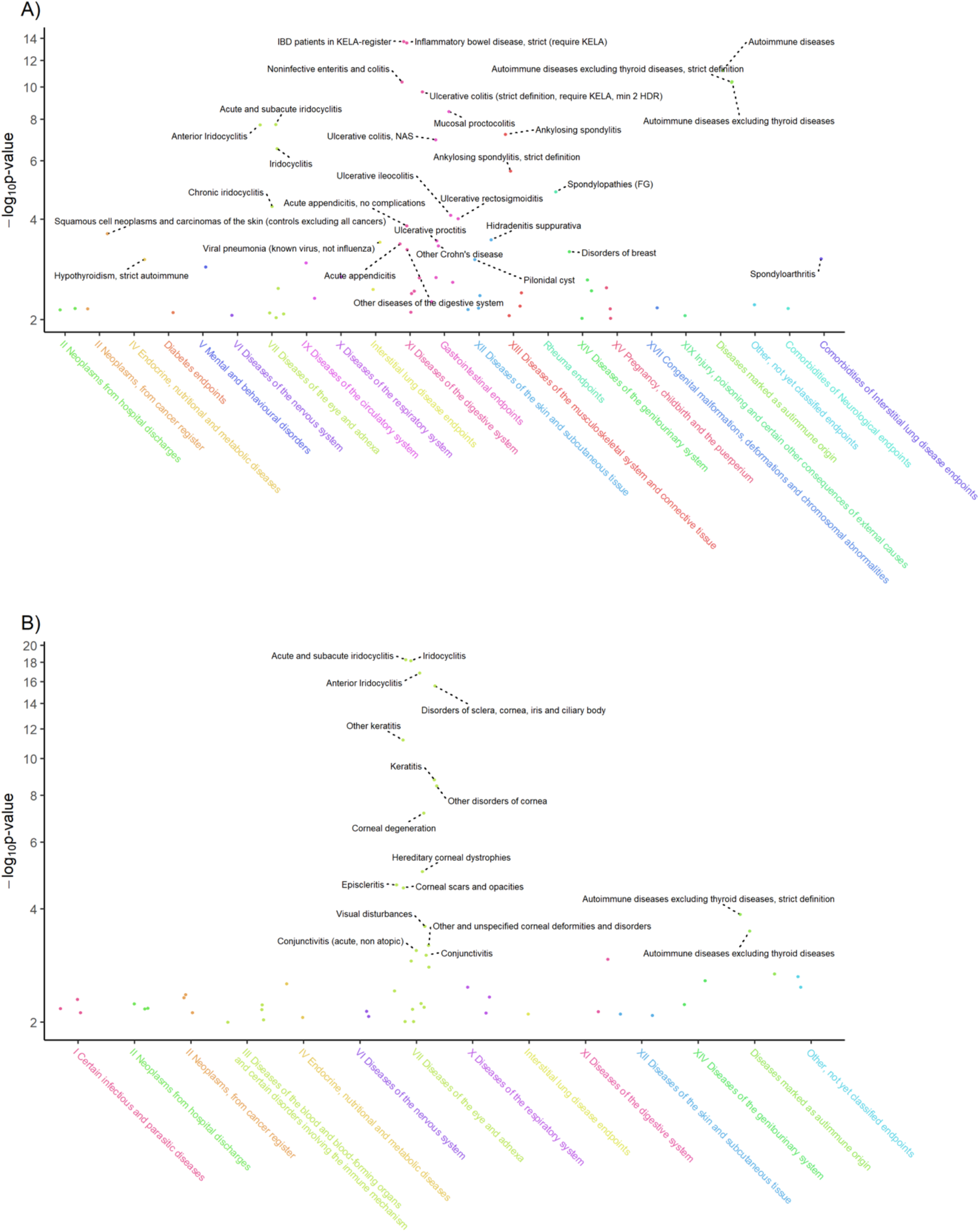
Associations of anterior uveitis associated loci across different phenotypes. Phenome-wide associations (*p* < 0.01) of two anterior uveitis associated loci, *INAVA* (panel A) and *NLRP3-OR2L13* (panel B), across FinnGen endpoints are shown. FinnGen endpoints are distributed along the x axis with general categories indicated below the plots, and the y axis shows the -log10 p values of the associations. SNP rs6701496 is associated with multiple immune-related phenotypes (panel A), and rs145759877 primarily with different subtypes of anterior uveitis and other eye diseases (panel B).

Next, we studied the expression and protein quantitative trait locus associations of the lead SNPs. We detected that the lead SNP rs6701496 near the *INAVA* gene (also known as *C1orf106*) was associated with the expression levels of the *C1orf106* gene in various tissues including whole blood, esophagus mucosa and different skin regions. In the evaluation of protein levels, the lead SNP rs572758335 at the *ERAP1* gene locus was associated with ERAP1 protein levels. No associations were detected between the other lead SNPs and mRNA or protein levels.

### Comparison to previous associations with anterior uveitis

We compared our lead SNPs to previously associated loci reported in GWASs of acute anterior uveitis (Supplementary Table S3).^16–18^ In addition to associations at the *HLA* region, associations at the *ERAP1* and *TNCR18* loci were replicated at genome-wide significance (*p* < 5 × 10^−8^).^16–18^ Additionally, previous associations were replicated at the *IL23R* (rs79755370, *p* = 9.46 × 10^−5^) and *HDAC2-AS2* (rs11153469, *p* = 0.005) loci.^17,18^ Although not reaching the level of significance, nominal association was identified at the *B3GNT2-TMEM17* (rs11153469, *p* = 0.018) locus previously associated with anterior uveitis.^17^ However, previously reported loci *SELENOV, CAVIN4* and *MAP6* were not associated with anterior uveitis in our study.^18^

### Genetic correlations

We analyzed the genetic correlations of anterior uveitis with 181 traits (Supplementary Table S5) utilizing meta-analysis results. The most significant genetic correlations (*p* < 0.01) are shown in Supplementary Figure S3. We identified a moderate genetic correlation between anterior uveitis and inflammatory bowel disease (*r*_*g*_=0.39, *p* = 8 × 10^−5^). Various nominal genetic correlations between anterior uveitis and other conditions were identified including Crohn’s disease (*r*_*g*_=0.41, *p* = 0.03), other serious medical condition/disability diagnosed by doctor (*r*_*g*_=0.30, *p* = 0.0009), other eye problems (*r*_*g*_=0.46, *p* = 0.0016) and overall health rating (*r*_*g*_=0.18, *p* = 0.0021). As participants of our meta-analysis were partly overlapping with those included in GWASs of the analyzed traits, we conducted additional genetic correlation analyses using anterior uveitis GWAS results from the FinnGen dataset only; this way we were able to utilize completely independent datasets in these analyses. We detected various nominal genetic correlations that were mostly comparable to the original results (Supplementary Table S5). These included genetic correlations between anterior uveitis and inflammatory bowel disease (*r*_*g*_=0.39, *p* = 0.0027) and other eye problems (*r*_*g*_=0.57, *p* = 0.0039).

We investigated the case overlap between anterior uveitis and inflammatory bowel disease in FinnGen. We detected that the case overlap was only 2.5% indicating that individuals diagnosed with anterior uveitis or inflammatory bowel disease largely represent distinct cases.

## DISCUSSION

In this genome-wide association study, we identified three novel anterior uveitis associated genomic regions with genome-wide significance: *INAVA, NLRP3* and *NOS2*. We also detected genome-wide significant associations at three previously reported loci, *ERAP1, TNRC18* and the *HLA* region, and replicated associations at *IL23R* and *HDAC2-AS2* loci. A moderate genetic correlation between anterior uveitis and inflammatory bowel disease was found. In phenome-wide association study, we further detected that three anterior uveitis associated loci (*ERAP1, INAVA* and *TNRC18*) were associated with other immunity-related phenotypes, such as ankylosing spondylitis and inflammatory bowel disease.

Here, the majority of the candidate genes is involved in the regulation of immune response. *INAVA* is crucial in optimal pattern recognition receptor induced signaling, as well as in the secretion of cytokines and the clearance of bacteria.^38^ It is recognized as a susceptibility locus for Crohn’s disease.^33^ In our phenome-wide association study, the lead SNP near the *INAVA* gene displays associations with lymphocyte percentage of white cells, inflammatory bowel disease and ulcerative colitis. This SNP is associated with the expression levels of the *INAVA* gene in various tissues, potentially implicating its role in systemic inflammatory conditions by affecting mRNA levels. These findings further highlight the potential shared mechanisms between anterior uveitis and other systemic inflammatory diseases.

In our study, we replicated the previously known association of *ERAP1* and anterior uveitis^17^, and detected that the lead SNP is associated with multiple autoimmune diseases including ankylosing spondylitis, Behçet’s disease and chronic inflammatory diseases. *ERAP1* has been associated with ankylosing spondylitis and Behçet disease also in previous studies.^39,40^ The effect of *ERAP1* in ankylosing spondylitis might result from alterations of the HLA-B27 peptidome, impacting the immunological features of HLA-B27.^40^ This observation is of interest as ankylosing spondylitis, *ERAP1* and HLA-B27 are associated also with anterior uveitis. Future research could provide further insights into the potential role of these factors in the pathogenesis of anterior uveitis.

Additionally, we replicated the known association of immunity regulator gene *TNRC18* and anterior uveitis.^20^ Previous research has demonstrated that loss of *TNRC18* leads to the activation of immunity-related genes such as *IL11* and *TLR4*.^37^ The lead SNP in this locus is associated with various immune-related diseases with the most significant association observed for inflammatory bowel disease.

Previous studies have identified *NOS2* as a known locus associated with ankylosing spondylitis and suggestively associated with acute anterior uveitis.^16^ Nitric oxide, largely produced by the NOS2 enzyme, is involved in immunity and inflammation.^34^ Excessive production of nitric oxide is linked to the pathogenesis of several inflammatory conditions such as inflammatory bowel disease, graft-versus-host-disease and asthma.^41–43^ We report for the first time a genome-wide significant association with anterior uveitis at this locus highlighting its importance in the inflammatory diseases.

Here, SNPs at the *OR2L13*/*NLRP3* locus appear to be more specifically associated with eye-related diseases. *NLRP3* pathogenic variants are known to induce monogenic autoinflammatory cryopyrin-associated periodic syndromes including Muckle-Wells syndrome in which uveitis is one possible form of ocular involvement.^44,45^ Previously, a missense mutation in the *NLRP3* gene has been discovered as the cause of keratoendotheliitis fugax hereditaria in Finnish population.^46^ Keratoendotheliitis fugax hereditaria manifests as an inflammatory assault affecting endothelium and posterior stroma of the cornea and it may be associated with a mild anterior chamber reaction.^46^ This further supports the role of *NLRP3* in inflammation of the anterior segment of the eye.

Further, we report a moderate genetic correlation between anterior uveitis and inflammatory bowel disease. In previous studies, ulcerative colitis and Crohn’s disease were associated with an increased risk of anterior uveitis, with a particularly heightened risk in Crohn’s disease. ^47–50^ Among individuals with inflammatory bowel disease, anterior uveitis has been documented in up to 17% of cases.^51^ In our study, the case overlap between anterior uveitis and inflammatory bowel disease was only 2.5% indicating that individuals diagnosed with anterior uveitis or inflammatory bowel disease largely represent distinct cases, with only a minimal overlap between the two conditions. This suggests the possibility of additional modifying factors determing the manner of immune-related reaction and the tissue site involved.

Strengths of our study are the large number of participants with a total of 12 205 cases and 917 145 controls, and the genetic composition of Finnish population. The bottlenecked genetic composition of Finnish population facilitates the detection of enriched genetic variants. Also, the FINNGEN study provides a rich resource to elucidate the specificity of the associated loci to a given outcome. While this study represents the largest GWAS meta-analysis specifically for anterior uveitis to date, some limitations must be acknowledged. The reliance on European cohorts limits the generalizability of the findings to other populations. However, this study deepens the understanding of biological mechanisms of anterior uveitis.

## CONCLUSIONS

In this genome-wide association study, we detected three novel loci associated with anterior uveitis. Previously reported associations at *HLA* region, *ERAP1* and *TNRC18* were replicated. In addition, a genetic correlation between anterior uveitis and inflammatory bowel disease was found. Our findings deepen our understanding of the genetic basis of anterior uveitis and the genetic connections between anterior uveitis and immune-related disorders, particularly inflammatory bowel disease, providing a foundation for further research and potential therapeutic interventions.

## Data Availability

Summary statistics will be made publicly available upon publication.

## ACKNOWLEDGEMENTS

We want to acknowledge the participants and investigators of FinnGen study. The FinnGen project is funded by two grants from Business Finland (HUS 4685/31/2016 and UH 4386/31/2016) and the following industry partners: AbbVie Inc., AstraZeneca UK Ltd, Biogen MA Inc., Bristol Myers Squibb (and Celgene Corporation & Celgene International II Sàrl), Genentech Inc., Merck Sharp & Dohme LCC, Pfizer Inc., GlaxoSmithKline Intellectual Property Development Ltd., Sanofi US Services Inc., Maze Therapeutics Inc., Janssen Biotech Inc, Novartis Pharma AG, and Boehringer Ingelheim International GmbH. Following biobanks are acknowledged for delivering biobank samples to FinnGen: Auria Biobank (www.auria.fi/biopankki), THL Biobank (www.thl.fi/biobank), Helsinki Biobank (www.helsinginbiopankki.fi), Biobank Borealis of Northern Finland (https://www.ppshp.fi/Tutkimus-ja-opetus/Biopankki/Pages/Biobank-Borealis-briefly-in-English.aspx), Finnish Clinical Biobank Tampere (www.tays.fi/en-US/Research_and_development/Finnish_Clinical_Biobank_Tampere), Biobank of Eastern Finland (www.ita-suomenbiopankki.fi/en), Central Finland Biobank (www.ksshp.fi/fi-FI/Potilaalle/Biopankki), Finnish Red Cross Blood Service Biobank (www.veripalvelu.fi/verenluovutus/biopankkitoiminta), Terveystalo Biobank (www.terveystalo.com/fi/Yritystietoa/Terveystalo-Biopankki/Biopankki/) and Arctic Biobank (https://www.oulu.fi/en/university/faculties-and-units/faculty-medicine/northern-finland-birth-cohorts-and-arctic-biobank). All Finnish Biobanks are members of BBMRI.fi infrastructure (www.bbmri.fi). Finnish Biobank Cooperative -FINBB (https://finbb.fi/) is the coordinator of BBMRI-ERIC operations in Finland. The Finnish biobank data can be accessed through the Fingenious® services (https://site.fingenious.fi/en/) managed by FINBB. We want to acknowledge the participants of the Estonian Biobank for their contributions. The Estonian Genome Center GWAS analyses were performed in the High Performance Computing Center, University of Tartu. The activities of the EstBB are regulated by the Human Genes Research Act. Individual level data analysis in EstBB was carried out under ethical approval 1.1-12/1020 from the Estonian Committee on Bioethics and Human Research (Estonian Ministry of Social Affairs). The work of the Estonian Genome Center, University of Tartu was funded by the Estonian Research Council Grant PRG1291, and University of Helsinki Grant VLTGI20638. This study has received funding from the Suomen Silmälääkäriyhdistys, the Silmäsäätiö Foundation, the Sokeain Ystävät Foundation, the Mary and Georg C. Ehrnrooth Foundation and the Evald ja Hilda Nissin säätiö Foundation. The funding organizations had no role in the design or conduct of this study. The authors have no conflict of interest.

