## Supplementary Text for "Genome-wide association study of anterior uveitis"

^3^ Estonia

^4^ Tays Eye Centre, Tampere University Hospital, Tampere, Finland

^5^ Research Unit of Population Health, Faculty of Medicine, University of Oulu, Finland

^6^ Biocenter Oulu, University of Oulu, Oulu, Finland

^7^ Finnish Institute for Health and Welfare (THL), Helsinki, Finland

^8^ Northern Finland Birth Cohorts, Arctic Biobank, Infrastructure for Population Studies, Faculty of Medicine, University of Oulu, Oulu, Finland

^9^ Institute for Molecular Medicine Finland, Helsinki Institute of Life Science, University of Helsinki, Helsinki, Finland

**SUPPLEMENTARY MATERIAL**

**Table of contents**

Supplementary Note. FinnGen DF10 ethics statement.....................................................................................2

Supplementary text: Detailed descriptions of study populations……………………………………………………………......4

Figure S1. Regional association plots of genome-wide significant anterior uveitis associated loci..................6

Figure S2. Genetic correlations of anterior uveitis………...……………………………………………………………………………9

Figure S3. Forest plot showing effect estimates of lead single nucleotide polymorphisms in each study population (FinnGen, Estonian Biobank and UK Biobank) and meta-analysis………………………………………….….10

Supplementary text: ESTBB consortium authors………………………………………………………………………………………11

Supplementary text: FinnGen consortium authors……………………………………………………………………………………12

**SUPPLEMENTARY NOTE: FinnGen DF10 ethics statement.**

Patients and control subjects in FinnGen provided informed consent for biobank research, based on the Finnish Biobank Act. Alternatively, separate research cohorts, collected prior the Finnish Biobank Act came into effect (in September 2013) and start of FinnGen (August 2017), were collected based on study-specific consents and later transferred to the Finnish biobanks after approval by Fimea (Finnish Medicines Agency), the National Supervisory Authority for Welfare and Health. Recruitment protocols followed the biobank protocols approved by Fimea. The Coordinating Ethics Committee of the Hospital District of Helsinki and Uusimaa (HUS) statement number for the FinnGen study is Nr HUS/990/2017.

The FinnGen study is approved by Finnish Institute for Health and Welfare (permit numbers: THL/2031/6.02.00/2017, THL/1101/5.05.00/2017, THL/341/6.02.00/2018, THL/2222/6.02.00/2018, THL/283/6.02.00/2019, THL/1721/5.05.00/2019 and THL/1524/5.05.00/2020), Digital and population data service agency (permit numbers: VRK43431/2017-3, VRK/6909/2018-3, VRK/4415/2019-3), the Social Insurance Institution (permit numbers: KELA 58/522/2017, KELA 131/522/2018, KELA 70/522/2019, KELA 98/522/2019, KELA 134/522/2019, KELA 138/522/2019, KELA 2/522/2020, KELA 16/522/2020), Findata permit numbers THL/2364/14.02/2020, THL/4055/14.06.00/2020, THL/3433/14.06.00/2020, THL/4432/14.06/2020, THL/5189/14.06/2020, THL/5894/14.06.00/2020, THL/6619/14.06.00/2020, THL/209/14.06.00/2021, THL/688/14.06.00/2021, THL/1284/14.06.00/2021, THL/1965/14.06.00/2021, THL/5546/14.02.00/2020, THL/2658/14.06.00/2021, THL/4235/14.06.00/2021, Statistics Finland (permit numbers: TK-53-1041-17 and TK/143/07.03.00/2020 (earlier TK-53-90-20) TK/1735/07.03.00/2021, TK/3112/07.03.00/2021) and Finnish Registry for Kidney Diseases permission/extract from the meeting minutes on 4th July 2019.

The Biobank Access Decisions for FinnGen samples and data utilized in FinnGen Data Freeze 10 include: THL Biobank BB2017_55, BB2017_111, BB2018_19, BB_2018_34, BB_2018_67, BB2018_71, BB2019_7, BB2019_8, BB2019_26, BB2020_1, BB2021_65, Finnish Red Cross Blood Service Biobank 7.12.2017, Helsinki Biobank HUS/359/2017, HUS/248/2020, HUS/150/2022 § 12, §13, §14, §15, §16, §17, §18, and §23, Auria Biobank AB17-5154 and amendment #1 (August 17 2020) and amendments BB_2021-0140, BB_2021-0156 (August 26 2021, Feb 2 2022), BB_2021-0169, BB_2021-0179, BB_2021-0161, AB20-5926 and amendment #1 (April 23 2020)and it´s modification (Sep 22 2021), Biobank Borealis of Northern Finland_2017_1013, 2021_5010, 2021_5018, 2021_5015, 2021_5023, 2021_5017, 2022_6001, Biobank of Eastern Finland 1186/2018 and amendment 22 § /2020, 53§/2021, 13§/2022, 14§/2022, 15§/2022, Finnish Clinical Biobank Tampere MH0004 and amendments (21.02.2020 & 06.10.2020), §8/2021, §9/2022, §10/2022, §12/2022, §20/2022, §21/2022, §22/2022, §23/2022, Central Finland Biobank 1-2017, and Terveystalo Biobank STB 2018001 and amendment 25th Aug 2020, Finnish Hematological Registry and Clinical Biobank decision 18th June 2021, Arctic biobank P0844: ARC_2021_1001.

**SUPPLEMENTARY TEXT: Detailed descriptions of study populations**

The FinnGen

The FinnGen study is a large-scale genomics initiative that has analyzed over 500,000 Finnish biobank samples and correlated genetic variation with health data to understand disease mechanisms and predispositions. The project is a collaboration between research organizations and biobanks within Finland and international industry partners.^1^ The present study included data from FinnGen Data Release R10.

The Estonian Biobank

The Estonian Biobank is a population-based biobank with 212,955 participants in the current data freeze (2023v4). All biobank participants have signed a broad informed consent form and information on ICD-10 codes is obtained via regular linking with the national Health Insurance Fund and other relevant databases, with majority of the electronic health records having been collected since 2004.^2^

All EstBB participants have been genotyped at the Core Genotyping Lab of the Institute of Genomics, University of Tartu, using Illumina Global Screening Array v1.0 and v2.0. Samples were genotyped, and PLINK format files were created using Illumina GenomeStudio v2.0.4. Individuals were excluded from the analysis if their call rate was <95% or if the sex defined based on heterozygosity of the X chromosome did not match the sex in phenotype data. Before imputation, variants were filtered by call rate <95%, Hardy–Weinberg equilibrium (HWE) P < 1 × 10−4 (autosomal variants only) and minor allele frequency <1%. Variant positions were updated to b37, and all variants were changed to be from TOP strand using GSAMD-24v1-0_20011747_A1-b37.strand.RefAlt.zip files from https://www.well.ox.ac.uk/~wrayner/strand/ webpage. Prephasing was done using Eagle v2.3 software38 (number of conditioning haplotypes Eagle2 uses when phasing each sample was set to:–Kpbwt=20000) and imputation was done using Beagle v.28Sep18.79339 with effective population size ne = 20,000. Population-specific imputation reference of 2,297 WGS samples was used.^3^

The activities of the EstBB are regulated by the Human Genes Research Act, which was adopted in 2000 specifically for the operations of EstBB. Individual level data analysis in EstBB was carried out under ethical approval nr 1.1-12/1020 from the Estonian Committee on Bioethics and Human Research (Estonian Ministry of Social Affairs), using data according to release application 3-10/GI/1915 from the Estonian Biobank.

The UK Biobank

The UK Biobank is a large-scale open database including a half million individuals with paired genetic and phenotype information that has been enormously valuable in studies of genetic etiology for common diseases and traits. The PanUKBB (<https://pan.ukbb.broadinstitute.org/>) project performed a pan-ancestry genetic analysis for 500 000 UK Biobank samples, with 40 to 69 years old adults as participants. The phenotypes were identified with registry data and questionnaires. The project is aiming to follow up participants for at least 30 years.^4^

1.	Kurki MI, Karjalainen J, Palta P, et al. FinnGen provides genetic insights from a well-phenotyped isolated population. *Nature*. 2023;613(7944):508-518. doi:10.1038/s41586-022-05473-8

2.	Leitsalu L, Haller T, Esko T, et al. Cohort Profile: Estonian Biobank of the Estonian Genome Center, University of Tartu. *Int J Epidemiol*. 2015;44(4):1137-1147. doi:10.1093/ije/dyt268

3.	Laisk T, Lepamets M, Koel M, Abner E, Estonian Biobank Research Team, Mägi R. Genome-wide association study identifies five risk loci for pernicious anemia. *Nat Commun*. 2021;12(1):3761. doi:10.1038/s41467-021-24051-6

4.	Sudlow C, Gallacher J, Allen N, et al. UK biobank: an open access resource for identifying the causes of a wide range of complex diseases of middle and old age. *PLoS Med*. 2015;12(3):e1001779. doi:10.1371/journal.pmed.1001779

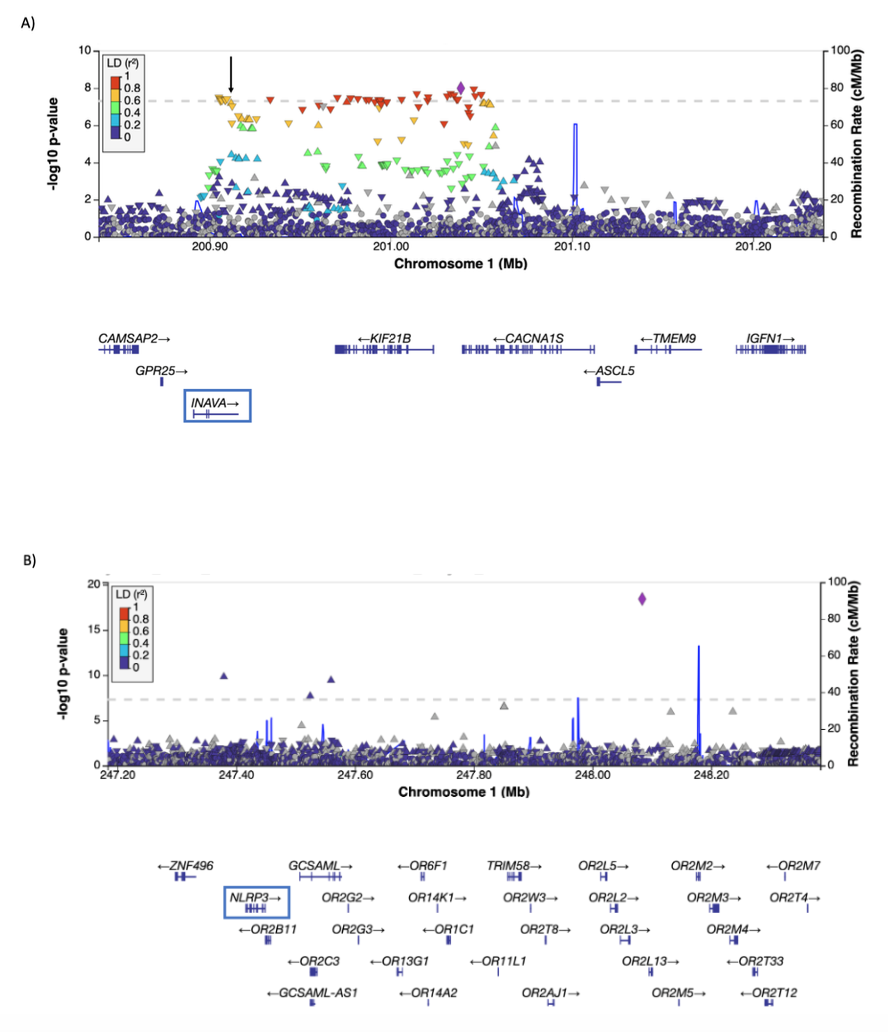

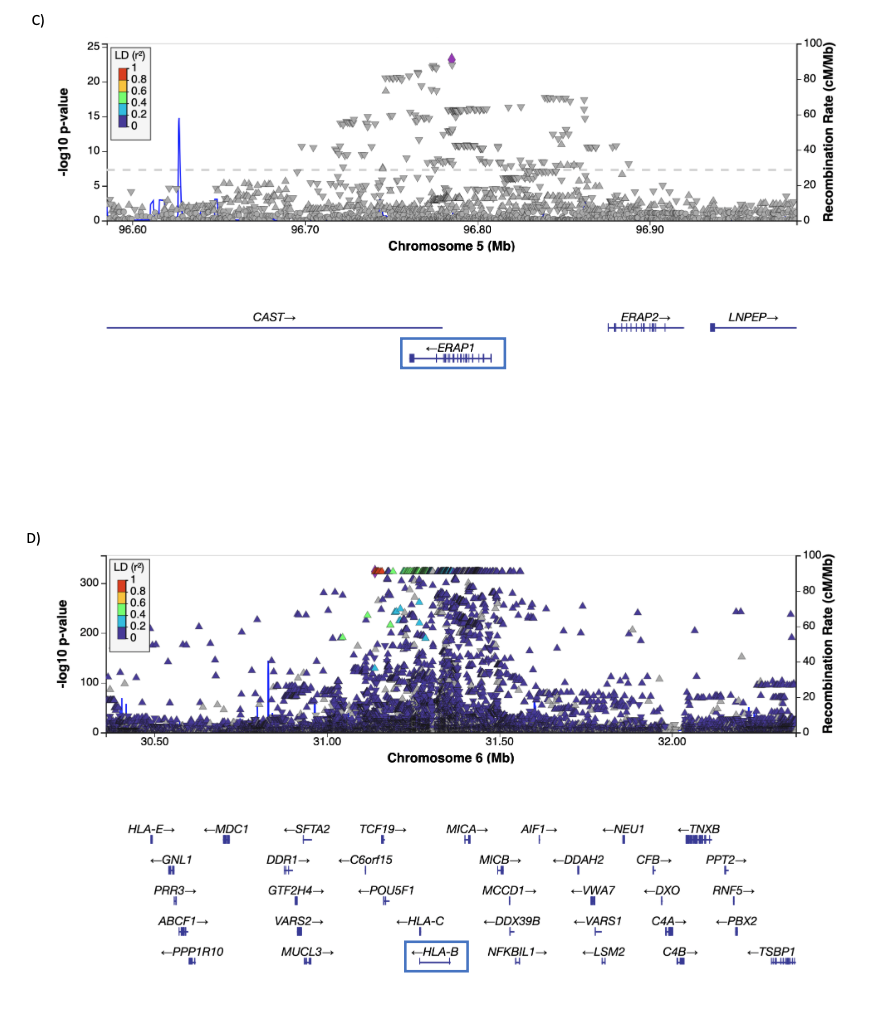

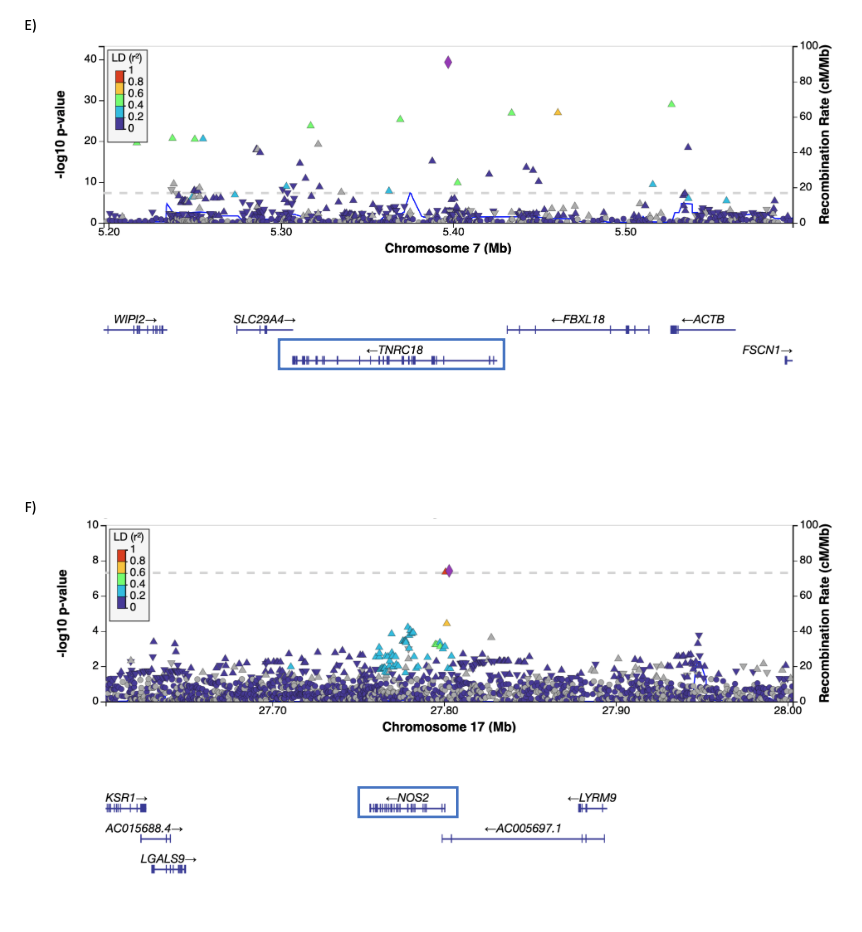

**Figure S1.** Regional association plots of genome-wide significant (*p* < 5 x 10^-8^) anterior uveitis associated loci A) *INAVA*, B) *NLRP3*, C) *ERAP1,* D) *HLA-B,* E) *TNRC18* and F) *NOS2*. Candidate genes are marked with a blue box. European LD reference panel is applied. The genome-wide significance threshold is marked with a dashed grey line. Missense SNP in linkage disequilibrium with the lead SNP near *INAVA* gene is marked with a black arrow.

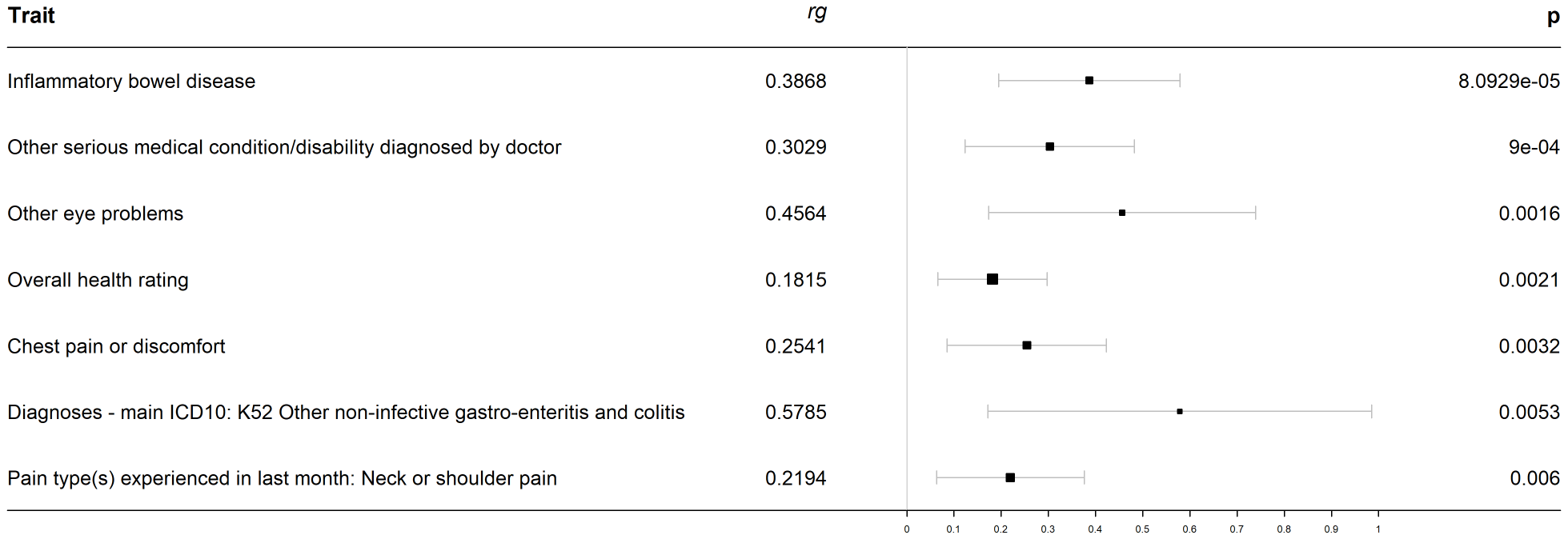

**Figure S2.** Genetic correlations of anterior uveitis with different traits using linkage disequilibrium score regression (LDSC). Table displays genetic correlations observed in the dataset with *p*-value <0.01.

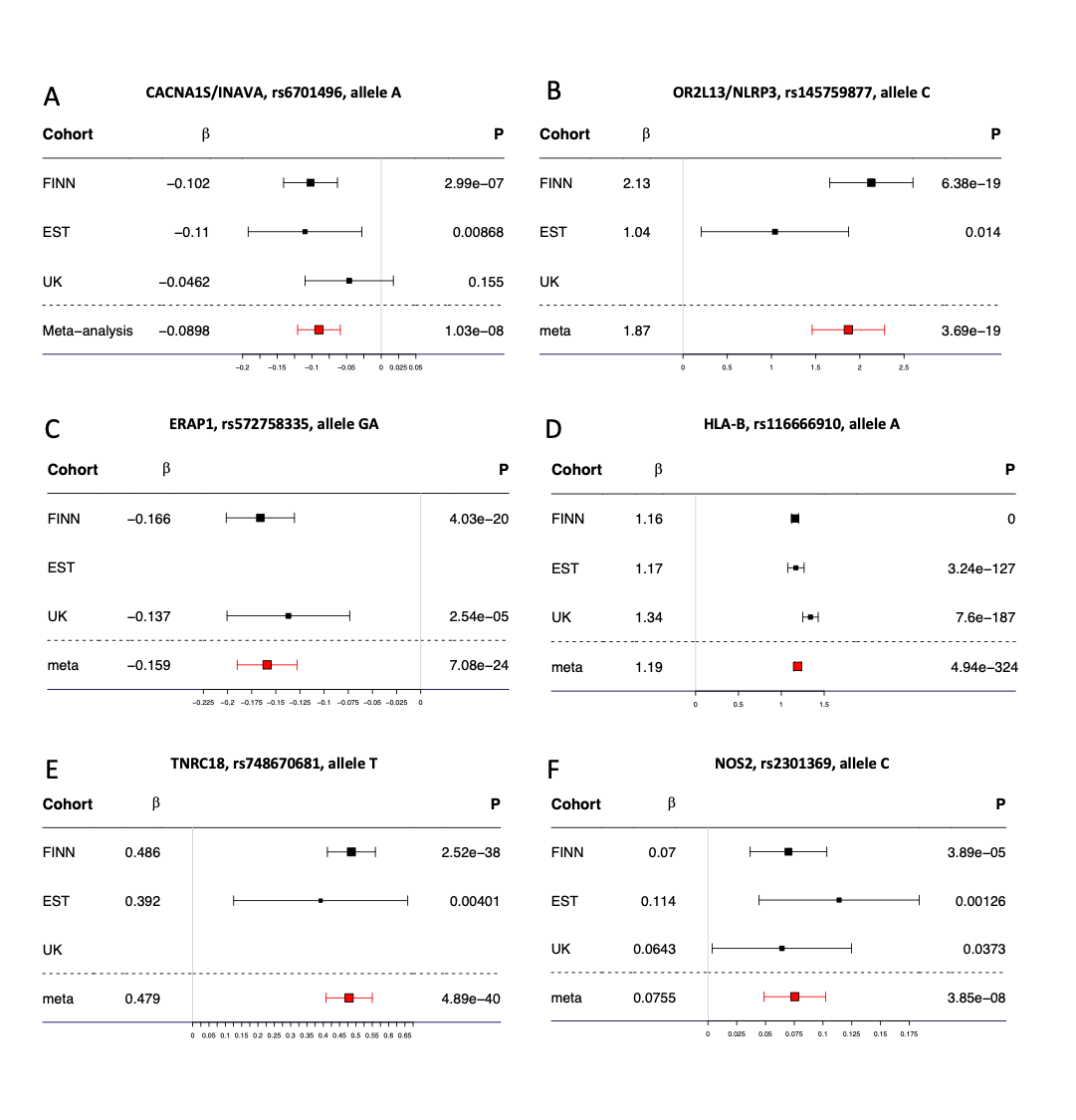

**Figure S3.** Forest plot showing effect estimates of lead single nucleotide polymorphisms in each study population (Finn, FinnGen; Est, Estonian Biobank; UK, UK Biobank) and meta-analysis. Whiskers are displaying the 95 % confidence interval.

**SUPPLEMENTARY TEXT: ESTBB consortium authors**

Data collection, genotyping, QC and imputation:

Andres Metspalu -

Lili Milani -

Reedik Mägi -

Mari Nelis -

Georgi Hudjashov -

Tõnu Esko -

**SUPPLEMENTARY TEXT: FinnGen consortium authors**

| Full Name | Affiliation | Role 1 | Role 2 |
| --- | --- | --- | --- |
| Aarno Palotie | Institute for Molecular Medicine Finland (FIMM), HiLIFE, University of Helsinki, Helsinki, Finland; Broad Institute of MIT and Harvard; Massachusetts General Hospital | Steering Committee | Steering Committee |
| Mark Daly | Institute for Molecular Medicine Finland (FIMM), HiLIFE, University of Helsinki, Helsinki, Finland; Broad Institute of MIT and Harvard; Massachusetts General Hospital | Steering Committee | Steering Committee |
| Bridget Riley-Gills | Abbvie, Chicago, IL, United States | Steering Committee | Pharmaceutical companies |
| Howard Jacob | Abbvie, Chicago, IL, United States | Steering Committee | Pharmaceutical companies |
| Dirk Paul | Astra Zeneca, Cambridge, United Kingdom | Steering Committee | Pharmaceutical companies |
| Slavé Petrovski | Astra Zeneca, Cambridge, United Kingdom | Steering Committee | Pharmaceutical companies |
| Chia-Yen Chen | Biogen, Cambridge, MA, United States | Steering Committee | Pharmaceutical companies |
| Sally John | Biogen, Cambridge, MA, United States | Steering Committee | Pharmaceutical companies |
| George Okafo | Boehringer Ingelheim, Ingelheim am Rhein, Germany | Steering Committee | Pharmaceutical companies |
| Robert Plenge | Bristol Myers Squibb, New York, NY, United States | Steering Committee | Pharmaceutical companies |
| Joseph Maranville | Bristol Myers Squibb, New York, NY, United States | Steering Committee | Pharmaceutical companies |
| Mark McCarthy | Genentech, San Francisco, CA, United States | Steering Committee | Pharmaceutical companies |
| Rion Pendergrass | Genentech, San Francisco, CA, United States | Steering Committee | Pharmaceutical companies |
| Margaret G. Ehm | GlaxoSmithKline, Collegeville, PA, United States | Steering Committee | Pharmaceutical companies |
| Kirsi Auro | GlaxoSmithKline, Espoo, Finland | Steering Committee | Pharmaceutical companies |
| Simonne Longerich | Merck, Kenilworth, NJ, United States | Steering Committee | Pharmaceutical companies |
| Anders Mälarstig | Pfizer, New York, NY, United States | Steering Committee | Pharmaceutical companies |
| Anna Vlahiotis | Pfizer, New York, NY, United States | Steering Committee | Pharmaceutical companies |
| Katherine Klinger | Translational Sciences, Sanofi R&D, Framingham, MA, USA | Steering Committee | Pharmaceutical companies |
| Clement Chatelain | Translational Sciences, Sanofi R&D, Framingham, MA, USA | Steering Committee | Pharmaceutical companies |
| Matthias Gossel | Translational Sciences, Sanofi R&D, Framingham, MA, USA | Steering Committee | Pharmaceutical companies |
| Karol Estrada | Maze Therapeutics, San Francisco, CA, United States | Steering Committee | Pharmaceutical companies |
| Robert Graham | Maze Therapeutics, San Francisco, CA, United States | Steering Committee | Pharmaceutical companies |
| Dawn Waterworth | Janssen Research & Development, LLC, Spring House, PA, United States | Steering Committee | Pharmaceutical companies |
| Chris O´Donnell | Novartis Institutes for BioMedical Research, Cambridge, MA, United States | Steering Committee | Pharmaceutical companies |
| Nicole Renaud | Novartis Institutes for BioMedical Research, Cambridge, MA, United States | Steering Committee | Pharmaceutical companies |
| Tomi P. Mäkelä | HiLIFE, University of Helsinki, Finland, Finland | Steering Committee | University of Helsinki & Biobanks |
| Jaakko Kaprio | Institute for Molecular Medicine Finland (FIMM), HiLIFE, University of Helsinki, Helsinki, Finland | Steering Committee | University of Helsinki & Biobanks |
| Minna Ruddock | Arctic biobank / University of Oulu | Steering Committee | University of Helsinki & Biobanks |
| Petri Virolainen | Auria Biobank / University of Turku / Hospital District of Southwest Finland, Turku, Finland | Steering Committee | University of Helsinki & Biobanks |
| Antti Hakanen | Auria Biobank / University of Turku / Hospital District of Southwest Finland, Turku, Finland | Steering Committee | University of Helsinki & Biobanks |
| Terhi Kilpi | THL Biobank / Finnish Institute for Health and Welfare (THL), Helsinki, Finland | Steering Committee | University of Helsinki & Biobanks |
| Markus Perola | THL Biobank / Finnish Institute for Health and Welfare (THL), Helsinki, Finland | Steering Committee | University of Helsinki & Biobanks |
| Jukka Partanen | Finnish Red Cross Blood Service / Finnish Hematology Registry and Clinical Biobank, Helsinki, Finland | Steering Committee | University of Helsinki & Biobanks |
| Taneli Raivio | Helsinki Biobank / Helsinki University and Hospital District of Helsinki and Uusimaa, Helsinki | Steering Committee | University of Helsinki & Biobanks |
| Jani Tikkanen | Northern Finland Biobank Borealis / University of Oulu / Northern Ostrobothnia Hospital District, Oulu, Finland | Steering Committee | University of Helsinki & Biobanks |
| Raisa Serpi | Northern Finland Biobank Borealis / University of Oulu / Northern Ostrobothnia Hospital District, Oulu, Finland | Steering Committee | University of Helsinki & Biobanks |
| Tarja Laitinen | Finnish Clinical Biobank Tampere / University of Tampere / Pirkanmaa Hospital District, Tampere, Finland | Steering Committee | University of Helsinki & Biobanks |
| Veli-Matti Kosma | Biobank of Eastern Finland / University of Eastern Finland / Northern Savo Hospital District, Kuopio, Finland | Steering Committee | University of Helsinki & Biobanks |
| Jari Laukkanen | Central Finland Biobank / University of Jyväskylä / Central Finland Health Care District, Jyväskylä, Finland | Steering Committee | University of Helsinki & Biobanks |
| Marco Hautalahti | FINBB - Finnish biobank cooperative | Steering Committee | University of Helsinki & Biobanks |
| Outi Tuovila | Business Finland, Helsinki, Finland | Steering Committee | Other Experts/ Non-Voting Members |
| Raimo Pakkanen | Business Finland, Helsinki, Finland | Steering Committee | Other Experts/ Non-Voting Members |
| Jeffrey Waring | Abbvie, Chicago, IL, United States | Scientific Committee | Pharmaceutical companies |
| Bridget Riley-Gillis | Abbvie, Chicago, IL, United States | Scientific Committee | Pharmaceutical companies |
| Fedik Rahimov | Abbvie, Chicago, IL, United States | Scientific Committee | Pharmaceutical companies |
| Ioanna Tachmazidou | Astra Zeneca, Cambridge, United Kingdom | Scientific Committee | Pharmaceutical companies |
| Chia-Yen Chen | Biogen, Cambridge, MA, United States | Scientific Committee | Pharmaceutical companies |
| Zhihao Ding | Boehringer Ingelheim, Ingelheim am Rhein, Germany | Scientific Committee | Pharmaceutical companies |
| Marc Jung | Boehringer Ingelheim, Ingelheim am Rhein, Germany | Scientific Committee | Pharmaceutical companies |
| Hanati Tuoken | Boehringer Ingelheim, Ingelheim am Rhein, Germany | Scientific Committee | Pharmaceutical companies |
| Shameek Biswas | Bristol Myers Squibb, New York, NY, United States | Scientific Committee | Pharmaceutical companies |
| Rion Pendergrass | Genentech, San Francisco, CA, United States | Scientific Committee | Pharmaceutical companies |
| Margaret G. Ehm | GlaxoSmithKline, Collegeville, PA, United States | Scientific Committee | Pharmaceutical companies |
| David Pulford | GlaxoSmithKline, Stevenage, United Kingdom | Scientific Committee | Pharmaceutical companies |
| Neha Raghavan | Merck, Kenilworth, NJ, United States | Scientific Committee | Pharmaceutical companies |
| Adriana Huertas-Vazquez | Merck, Kenilworth, NJ, United States | Scientific Committee | Pharmaceutical companies |
| Jae-Hoon Sul | Merck, Kenilworth, NJ, United States | Scientific Committee | Pharmaceutical companies |
| Anders Mälarstig | Pfizer, New York, NY, United States | Scientific Committee | Pharmaceutical companies |
| Xinli Hu | Pfizer, New York, NY, United States | Scientific Committee | Pharmaceutical companies |
| Åsa Hedman | Pfizer, New York, NY, United States | Scientific Committee | Pharmaceutical companies |
| Katherine Klinger | Translational Sciences, Sanofi R&D, Framingham, MA, USA | Scientific Committee | Pharmaceutical companies |
| Robert Graham | Maze Therapeutics, San Francisco, CA, United States | Scientific Committee | Pharmaceutical companies |
| Dawn Waterworth | Janssen Research & Development, LLC, Spring House, PA, United States | Scientific Committee | Pharmaceutical companies |
| Nicole Renaud | Novartis Institutes for BioMedical Research, Cambridge, MA, United States | Scientific Committee | Pharmaceutical companies |
| Ma´en Obeidat | Novartis Institutes for BioMedical Research, Cambridge, MA, United States | Scientific Committee | Pharmaceutical companies |
| Jonathan Chung | Novartis Institutes for BioMedical Research, Cambridge, MA, United States | Scientific Committee | Pharmaceutical companies |
| Jonas Zierer | Novartis Institutes for BioMedical Research, Cambridge, MA, United States | Scientific Committee | Pharmaceutical companies |
| Mari Niemi | Novartis Institutes for BioMedical Research, Cambridge, MA, United States | Scientific Committee | Pharmaceutical companies |
| Samuli Ripatti | Institute for Molecular Medicine Finland (FIMM), HiLIFE, University of Helsinki, Helsinki, Finland | Scientific Committee | University of Helsinki & Biobanks |
| Johanna Schleutker | Auria Biobank / Univ. of Turku / Hospital District of Southwest Finland, Turku, Finland | Scientific Committee | University of Helsinki & Biobanks |
| Markus Perola | THL Biobank / Finnish Institute for Health and Welfare (THL), Helsinki, Finland | Scientific Committee | University of Helsinki & Biobanks |
| Mikko Arvas | Finnish Red Cross Blood Service / Finnish Hematology Registry and Clinical Biobank, Helsinki, Finland | Scientific Committee | University of Helsinki & Biobanks |
| Olli Carpén | Helsinki Biobank / Helsinki University and Hospital District of Helsinki and Uusimaa, Helsinki | Scientific Committee | University of Helsinki & Biobanks |
| Reetta Hinttala | Northern Finland Biobank Borealis / University of Oulu / Northern Ostrobothnia Hospital District, Oulu, Finland | Scientific Committee | University of Helsinki & Biobanks |
| Johannes Kettunen | Northern Finland Biobank Borealis / University of Oulu / Northern Ostrobothnia Hospital District, Oulu, Finland | Scientific Committee | University of Helsinki & Biobanks |
| Arto Mannermaa | Biobank of Eastern Finland / University of Eastern Finland / Northern Savo Hospital District, Kuopio, Finland | Scientific Committee | University of Helsinki & Biobanks |
| Katriina Aalto-Setälä | Faculty of Medicine and Health Technology, Tampere University, Tampere, Finland | Scientific Committee | University of Helsinki & Biobanks |
| Mika Kähönen | Finnish Clinical Biobank Tampere / University of Tampere / Pirkanmaa Hospital District, Tampere, Finland | Scientific Committee | University of Helsinki & Biobanks |
| Jari Laukkanen | Central Finland Biobank / University of Jyväskylä / Central Finland Health Care District, Jyväskylä, Finland | Scientific Committee | University of Helsinki & Biobanks |
| Johanna Mäkelä | FINBB - Finnish biobank cooperative | Scientific Committee | University of Helsinki & Biobanks |
| Reetta Kälviäinen | Northern Savo Hospital District, Kuopio, Finland | Clinical Groups | Neurology Group |
| Valtteri Julkunen | Northern Savo Hospital District, Kuopio, Finland | Clinical Groups | Neurology Group |
| Hilkka Soininen | Northern Savo Hospital District, Kuopio, Finland | Clinical Groups | Neurology Group |
| Anne Remes | Northern Ostrobothnia Hospital District, Oulu, Finland | Clinical Groups | Neurology Group |
| Mikko Hiltunen | University of Eastern Finland, Kuopio, Finland | Clinical Groups | Neurology Group |
| Jukka Peltola | Pirkanmaa Hospital District, Tampere, Finland | Clinical Groups | Neurology Group |
| Minna Raivio | Hospital District of Helsinki and Uusimaa, Helsinki, Finland | Clinical Groups | Neurology Group |
| Pentti Tienari | Hospital District of Helsinki and Uusimaa, Helsinki, Finland | Clinical Groups | Neurology Group |
| Juha Rinne | Hospital District of Southwest Finland, Turku, Finland | Clinical Groups | Neurology Group |
| Roosa Kallionpää | Hospital District of Southwest Finland, Turku, Finland | Clinical Groups | Neurology Group |
| Juulia Partanen | Institute for Molecular Medicine Finland, HiLIFE, University of Helsinki, Finland | Clinical Groups | Neurology Group |
| Adam Ziemann | Abbvie, Chicago, IL, United States | Clinical Groups | Neurology Group |
| Nizar Smaoui | Abbvie, Chicago, IL, United States | Clinical Groups | Neurology Group |
| Anne Lehtonen | Abbvie, Chicago, IL, United States | Clinical Groups | Neurology Group |
| Susan Eaton | Biogen, Cambridge, MA, United States | Clinical Groups | Neurology Group |
| Heiko Runz | Biogen, Cambridge, MA, United States | Clinical Groups | Neurology Group |
| Sanni Lahdenperä | Biogen, Cambridge, MA, United States | Clinical Groups | Neurology Group |
| Shameek Biswas | Bristol Myers Squibb, New York, NY, United States | Clinical Groups | Neurology Group |
| Natalie Bowers | Genentech, San Francisco, CA, United States | Clinical Groups | Neurology Group |
| Edmond Teng | Genentech, San Francisco, CA, United States | Clinical Groups | Neurology Group |
| Rion Pendergrass | Genentech, San Francisco, CA, United States | Clinical Groups | Neurology Group |
| Fanli Xu | GlaxoSmithKline, Brentford, United Kingdom | Clinical Groups | Neurology Group |
| David Pulford | GlaxoSmithKline, Stevenage, United Kingdom | Clinical Groups | Neurology Group |
| Kirsi Auro | GlaxoSmithKline, Espoo, Finland | Clinical Groups | Neurology Group |
| Laura Addis | GlaxoSmithKline, Brentford, United Kingdom | Clinical Groups | Neurology Group |
| John Eicher | GlaxoSmithKline, Brentford, United Kingdom | Clinical Groups | Neurology Group |
| Qingqin S Li | Janssen Research & Development, LLC, Titusville, NJ 08560, United States | Clinical Groups | Neurology Group |
| Karen He | Janssen Research & Development, LLC, Spring House, PA, United States | Clinical Groups | Neurology Group |
| Ekaterina Khramtsova | Janssen Research & Development, LLC, Spring House, PA, United States | Clinical Groups | Neurology Group |
| Neha Raghavan | Merck, Kenilworth, NJ, United States | Clinical Groups | Neurology Group |
| Martti Färkkilä | Hospital District of Helsinki and Uusimaa, Helsinki, Finland | Clinical Groups | Gastroenterology Group |
| Jukka Koskela | Hospital District of Helsinki and Uusimaa, Helsinki, Finland | Clinical Groups | Gastroenterology Group |
| Sampsa Pikkarainen | Hospital District of Helsinki and Uusimaa, Helsinki, Finland | Clinical Groups | Gastroenterology Group |
| Airi Jussila | Pirkanmaa Hospital District, Tampere, Finland | Clinical Groups | Gastroenterology Group |
| Katri Kaukinen | Pirkanmaa Hospital District, Tampere, Finland | Clinical Groups | Gastroenterology Group |
| Timo Blomster | Northern Ostrobothnia Hospital District, Oulu, Finland | Clinical Groups | Gastroenterology Group |
| Mikko Kiviniemi | Northern Savo Hospital District, Kuopio, Finland | Clinical Groups | Gastroenterology Group |
| Markku Voutilainen | Hospital District of Southwest Finland, Turku, Finland | Clinical Groups | Gastroenterology Group |
| Mark Daly | Institute for Molecular Medicine, Finland (FIMM), HiLIFE, University of Helsinki, Helsinki, Finland; Broad Institute of MIT and Harvard; Massachusetts General Hospital | Clinical Groups | Gastroenterology Group |
| Jeffrey Waring | Abbvie, Chicago, IL, United States | Clinical Groups | Gastroenterology Group |
| Nizar Smaoui | Abbvie, Chicago, IL, United States | Clinical Groups | Gastroenterology Group |
| Fedik Rahimov | Abbvie, Chicago, IL, United States | Clinical Groups | Gastroenterology Group |
| Anne Lehtonen | Abbvie, Chicago, IL, United States | Clinical Groups | Gastroenterology Group |
| Tim Lu | Genentech, San Francisco, CA, United States | Clinical Groups | Gastroenterology Group |
| Natalie Bowers | Genentech, San Francisco, CA, United States | Clinical Groups | Gastroenterology Group |
| Rion Pendergrass | Genentech, San Francisco, CA, United States | Clinical Groups | Gastroenterology Group |
| Linda McCarthy | GlaxoSmithKline, Brentford, United Kingdom | Clinical Groups | Gastroenterology Group |
| Amy Hart | Janssen Research & Development, LLC, Spring House, PA, United States | Clinical Groups | Gastroenterology Group |
| Meijian Guan | Janssen Research & Development, LLC, Spring House, PA, United States | Clinical Groups | Gastroenterology Group |
| Jason Miller | Merck, Kenilworth, NJ, United States | Clinical Groups | Gastroenterology Group |
| Kirsi Kalpala | Pfizer, New York, NY, United States | Clinical Groups | Gastroenterology Group |
| Melissa Miller | Pfizer, New York, NY, United States | Clinical Groups | Gastroenterology Group |
| Xinli Hu | Pfizer, New York, NY, United States | Clinical Groups | Gastroenterology Group |
| Kari Eklund | Hospital District of Helsinki and Uusimaa, Helsinki, Finland | Clinical Groups | Rheumatology Group |
| Antti Palomäki | Hospital District of Southwest Finland, Turku, Finland | Clinical Groups | Rheumatology Group |
| Pia Isomäki | Pirkanmaa Hospital District, Tampere, Finland | Clinical Groups | Rheumatology Group |
| Laura Pirilä | Hospital District of Southwest Finland, Turku, Finland | Clinical Groups | Rheumatology Group |
| Oili Kaipiainen-Seppänen | Northern Savo Hospital District, Kuopio, Finland | Clinical Groups | Rheumatology Group |
| Johanna Huhtakangas | Northern Ostrobothnia Hospital District, Oulu, Finland | Clinical Groups | Rheumatology Group |
| Nina Mars | Institute for Molecular Medicine Finland (FIMM), HiLIFE, University of Helsinki, Helsinki, Finland | Clinical Groups | Rheumatology Group |
| Jeffrey Waring | Abbvie, Chicago, IL, United States | Clinical Groups | Rheumatology Group |
| Fedik Rahimov | Abbvie, Chicago, IL, United States | Clinical Groups | Rheumatology Group |
| Apinya Lertratanakul | Abbvie, Chicago, IL, United States | Clinical Groups | Rheumatology Group |
| Nizar Smaoui | Abbvie, Chicago, IL, United States | Clinical Groups | Rheumatology Group |
| Anne Lehtonen | Abbvie, Chicago, IL, United States | Clinical Groups | Rheumatology Group |
| Coralie Viollet | AstraZeneca, Cambridge, United Kingdom | Clinical Groups | Rheumatology Group |
| Marla Hochfeld | Bristol Myers Squibb, New York, NY, United States | Clinical Groups | Rheumatology Group |
| Natalie Bowers | Genentech, San Francisco, CA, United States | Clinical Groups | Rheumatology Group |
| Rion Pendergrass | Genentech, San Francisco, CA, United States | Clinical Groups | Rheumatology Group |
| Jorge Esparza Gordillo | GlaxoSmithKline, Brentford, United Kingdom | Clinical Groups | Rheumatology Group |
| Kirsi Auro | GlaxoSmithKline, Espoo, Finland | Clinical Groups | Rheumatology Group |
| Dawn Waterworth | Janssen Research & Development, LLC, Spring House, PA, United States | Clinical Groups | Rheumatology Group |
| Fabiana Farias | Merck, Kenilworth, NJ, United States | Clinical Groups | Rheumatology Group |
| Kirsi Kalpala | Pfizer, New York, NY, United States | Clinical Groups | Rheumatology Group |
| Nan Bing | Pfizer, New York, NY, United States | Clinical Groups | Rheumatology Group |
| Xinli Hu | Pfizer, New York, NY, United States | Clinical Groups | Rheumatology Group |
| Tarja Laitinen | Pirkanmaa Hospital District, Tampere, Finland | Clinical Groups | Pulmonology Group |
| Margit Pelkonen | Northern Savo Hospital District, Kuopio, Finland | Clinical Groups | Pulmonology Group |
| Paula Kauppi | Hospital District of Helsinki and Uusimaa, Helsinki, Finland | Clinical Groups | Pulmonology Group |
| Hannu Kankaanranta | University of Gothenburg, Gothenburg, Sweden/ Seinäjoki Central Hospital, Seinäjoki, Finland/ Tampere University, Tampere, Finland | Clinical Groups | Pulmonology Group |
| Terttu Harju | Northern Ostrobothnia Hospital District, Oulu, Finland | Clinical Groups | Pulmonology Group |
| Riitta Lahesmaa | Hospital District of Southwest Finland, Turku, Finland | Clinical Groups | Pulmonology Group |
| Nizar Smaoui | Abbvie, Chicago, IL, United States | Clinical Groups | Pulmonology Group |
| Coralie Viollet | AstraZeneca, Cambridge, United Kingdom | Clinical Groups | Pulmonology Group |
| Susan Eaton | Biogen, Cambridge, MA, United States | Clinical Groups | Pulmonology Group |
| Hubert Chen | Genentech, San Francisco, CA, United States | Clinical Groups | Pulmonology Group |
| Rion Pendergrass | Genentech, San Francisco, CA, United States | Clinical Groups | Pulmonology Group |
| Natalie Bowers | Genentech, San Francisco, CA, United States | Clinical Groups | Pulmonology Group |
| Joanna Betts | GlaxoSmithKline, Brentford, United Kingdom | Clinical Groups | Pulmonology Group |
| Kirsi Auro | GlaxoSmithKline, Espoo, Finland | Clinical Groups | Pulmonology Group |
| Rajashree Mishra | GlaxoSmithKline, Brentford, United Kingdom | Clinical Groups | Pulmonology Group |
| Majd Mouded | Novartis, Basel, Switzerland | Clinical Groups | Pulmonology Group |
| Debby Ngo | Novartis, Basel, Switzerland | Clinical Groups | Pulmonology Group |
| Teemu Niiranen | Finnish Institute for Health and Welfare (THL), Helsinki, Finland | Clinical Groups | Cardiometabolic Diseases Group |
| Felix Vaura | Finnish Institute for Health and Welfare (THL), Helsinki, Finland | Clinical Groups | Cardiometabolic Diseases Group |
| Veikko Salomaa | Finnish Institute for Health and Welfare (THL), Helsinki, Finland | Clinical Groups | Cardiometabolic Diseases Group |
| Kaj Metsärinne | Hospital District of Southwest Finland, Turku, Finland | Clinical Groups | Cardiometabolic Diseases Group |
| Jenni Aittokallio | Hospital District of Southwest Finland, Turku, Finland | Clinical Groups | Cardiometabolic Diseases Group |
| Mika Kähönen | Pirkanmaa Hospital District, Tampere, Finland | Clinical Groups | Cardiometabolic Diseases Group |
| Jussi Hernesniemi | Pirkanmaa Hospital District, Tampere, Finland | Clinical Groups | Cardiometabolic Diseases Group |
| Daniel Gordin | Hospital District of Helsinki and Uusimaa, Helsinki, Finland | Clinical Groups | Cardiometabolic Diseases Group |
| Juha Sinisalo | Hospital District of Helsinki and Uusimaa, Helsinki, Finland | Clinical Groups | Cardiometabolic Diseases Group |
| Marja-Riitta Taskinen | Hospital District of Helsinki and Uusimaa, Helsinki, Finland | Clinical Groups | Cardiometabolic Diseases Group |
| Tiinamaija Tuomi | Hospital District of Helsinki and Uusimaa, Helsinki, Finland | Clinical Groups | Cardiometabolic Diseases Group |
| Timo Hiltunen | Hospital District of Helsinki and Uusimaa, Helsinki, Finland | Clinical Groups | Cardiometabolic Diseases Group |
| Jari Laukkanen | Central Finland Health Care District, Jyväskylä, Finland | Clinical Groups | Cardiometabolic Diseases Group |
| Amanda Elliott | Institute for Molecular Medicine Finland (FIMM), HiLIFE, University of Helsinki, Helsinki, Finland; Broad Institute, Cambridge, MA, USA and Massachusetts General Hospital, Boston, MA, USA | Clinical Groups | Cardiometabolic Diseases Group |
| Mary Pat Reeve | Institute for Molecular Medicine Finland (FIMM), HiLIFE, University of Helsinki, Helsinki, Finland | Clinical Groups | Cardiometabolic Diseases Group |
| Sanni Ruotsalainen | Institute for Molecular Medicine Finland (FIMM), HiLIFE, University of Helsinki, Helsinki, Finland | Clinical Groups | Cardiometabolic Diseases Group |
| Dirk Paul | Astra Zeneca, Cambridge, United Kingdom | Clinical Groups | Cardiometabolic Diseases Group |
| Natalie Bowers | Genentech, San Francisco, CA, United States | Clinical Groups | Cardiometabolic Diseases Group |
| Rion Pendergrass | Genentech, San Francisco, CA, United States | Clinical Groups | Cardiometabolic Diseases Group |
| Audrey Chu | GlaxoSmithKline, Brentford, United Kingdom | Clinical Groups | Cardiometabolic Diseases Group |
| Kirsi Auro | GlaxoSmithKline, Espoo, Finland | Clinical Groups | Cardiometabolic Diseases Group |
| Dermot Reilly | Janssen Research & Development, LLC, Boston, MA, United States | Clinical Groups | Cardiometabolic Diseases Group |
| Mike Mendelson | Novartis, Boston, MA, United States | Clinical Groups | Cardiometabolic Diseases Group |
| Jaakko Parkkinen | Pfizer, New York, NY, United States | Clinical Groups | Cardiometabolic Diseases Group |
| Melissa Miller | Pfizer, New York, NY, United States | Clinical Groups | Cardiometabolic Diseases Group |
| Tuomo Meretoja | Hospital District of Helsinki and Uusimaa, Helsinki, Finland | Clinical Groups | Oncology Group |
| Heikki Joensuu | Hospital District of Helsinki and Uusimaa, Helsinki, Finland | Clinical Groups | Oncology Group |
| Olli Carpén | Hospital District of Helsinki and Uusimaa, Helsinki, Finland | Clinical Groups | Oncology Group |
| Johanna Mattson | Hospital District of Helsinki and Uusimaa, Helsinki, Finland | Clinical Groups | Oncology Group |
| Eveliina Salminen | Hospital District of Helsinki and Uusimaa, Helsinki, Finland | Clinical Groups | Oncology Group |
| Annika Auranen | Pirkanmaa Hospital District , Tampere, Finland | Clinical Groups | Oncology Group |
| Peeter Karihtala | Northern Ostrobothnia Hospital District, Oulu, Finland | Clinical Groups | Oncology Group |
| Päivi Auvinen | Northern Savo Hospital District, Kuopio, Finland | Clinical Groups | Oncology Group |
| Klaus Elenius | Hospital District of Southwest Finland, Turku, Finland | Clinical Groups | Oncology Group |
| Johanna Schleutker | Hospital District of Southwest Finland, Turku, Finland | Clinical Groups | Oncology Group |
| Esa Pitkänen | Institute for Molecular Medicine Finland (FIMM), HiLIFE, University of Helsinki, Helsinki, Finland | Clinical Groups | Oncology Group |
| Nina Mars | Institute for Molecular Medicine Finland (FIMM), HiLIFE, University of Helsinki, Helsinki, Finland | Clinical Groups | Oncology Group |
| Mark Daly | Institute for Molecular Medicine Finland (FIMM), HiLIFE, University of Helsinki, Helsinki, Finland; Broad Institute of MIT and Harvard; Massachusetts General Hospital | Clinical Groups | Oncology Group |
| Relja Popovic | Abbvie, Chicago, IL, United States | Clinical Groups | Oncology Group |
| Jeffrey Waring | Abbvie, Chicago, IL, United States | Clinical Groups | Oncology Group |
| Bridget Riley-Gillis | Abbvie, Chicago, IL, United States | Clinical Groups | Oncology Group |
| Anne Lehtonen | Abbvie, Chicago, IL, United States | Clinical Groups | Oncology Group |
| Margarete Fabre | AstraZeneca, Cambridge, United Kingdom | Clinical Groups | Oncology Group |
| Jennifer Schutzman | Genentech, San Francisco, CA, United States | Clinical Groups | Oncology Group |
| Natalie Bowers | Genentech, San Francisco, CA, United States | Clinical Groups | Oncology Group |
| Rion Pendergrass | Genentech, San Francisco, CA, United States | Clinical Groups | Oncology Group |
| Diptee Kulkarni | GlaxoSmithKline, Brentford, United Kingdom | Clinical Groups | Oncology Group |
| Kirsi Auro | GlaxoSmithKline, Espoo, Finland | Clinical Groups | Oncology Group |
| Alessandro Porello | Janssen Research & Development, LLC, Spring House, PA, United States | Clinical Groups | Oncology Group |
| Andrey Loboda | Merck, Kenilworth, NJ, United States | Clinical Groups | Oncology Group |
| Heli Lehtonen | Pfizer, New York, NY, United States | Clinical Groups | Oncology Group |
| Stefan McDonough | Pfizer, New York, NY, United States | Clinical Groups | Oncology Group |
| Sauli Vuoti | Janssen-Cilag Oy, Espoo, Finland | Clinical Groups | Oncology Group |
| Kai Kaarniranta | Northern Savo Hospital District, Kuopio, Finland; Department of Molecular Genetics, University of Lodz, Lodz, Poland | Clinical Groups | Opthalmology Group |
| Joni A Turunen | Helsinki University Hospital and University of Helsinki, Helsinki, Finland; Eye Genetics Group, Folkhälsan Research Center, Helsinki, Finland | Clinical Groups | Opthalmology Group |
| Terhi Ollila | Hospital District of Helsinki and Uusimaa, Helsinki, Finland | Clinical Groups | Opthalmology Group |
| Hannu Uusitalo | Pirkanmaa Hospital District, Tampere, Finland | Clinical Groups | Opthalmology Group |
| Juha Karjalainen | Institute for Molecular Medicine Finland (FIMM), HiLIFE, University of Helsinki, Helsinki, Finland | Clinical Groups | Opthalmology Group |
| Esa Pitkänen | Institute for Molecular Medicine Finland (FIMM), HiLIFE, University of Helsinki, Helsinki, Finland | Clinical Groups | Opthalmology Group |
| Mengzhen Liu | Abbvie, Chicago, IL, United States | Clinical Groups | Opthalmology Group |
| Heiko Runz | Biogen, Cambridge, MA, United States | Clinical Groups | Opthalmology Group |
| Stephanie Loomis | Biogen, Cambridge, MA, United States | Clinical Groups | Opthalmology Group |
| Erich Strauss | Genentech, San Francisco, CA, United States | Clinical Groups | Opthalmology Group |
| Natalie Bowers | Genentech, San Francisco, CA, United States | Clinical Groups | Opthalmology Group |
| Hao Chen | Genentech, San Francisco, CA, United States | Clinical Groups | Opthalmology Group |
| Rion Pendergrass | Genentech, San Francisco, CA, United States | Clinical Groups | Opthalmology Group |
| Kaisa Tasanen | Northern Ostrobothnia Hospital District, Oulu, Finland | Clinical Groups | Dermatology Group |
| Laura Huilaja | Northern Ostrobothnia Hospital District, Oulu, Finland | Clinical Groups | Dermatology Group |
| Katariina Hannula-Jouppi | Hospital District of Helsinki and Uusimaa, Helsinki, Finland | Clinical Groups | Dermatology Group |
| Teea Salmi | Pirkanmaa Hospital District, Tampere, Finland | Clinical Groups | Dermatology Group |
| Sirkku Peltonen | Hospital District of Southwest Finland, Turku, Finland | Clinical Groups | Dermatology Group |
| Leena Koulu | Hospital District of Southwest Finland, Turku, Finland | Clinical Groups | Dermatology Group |
| Nizar Smaoui | Abbvie, Chicago, IL, United States | Clinical Groups | Dermatology Group |
| Fedik Rahimov | Abbvie, Chicago, IL, United States | Clinical Groups | Dermatology Group |
| Anne Lehtonen | Abbvie, Chicago, IL, United States | Clinical Groups | Dermatology Group |
| David Choy | Genentech, San Francisco, CA, United States | Clinical Groups | Dermatology Group |
| Rion Pendergrass | Genentech, San Francisco, CA, United States | Clinical Groups | Dermatology Group |
| Dawn Waterworth | Janssen Research & Development, LLC, Spring House, PA, United States | Clinical Groups | Dermatology Group |
| Kirsi Kalpala | Pfizer, New York, NY, United States | Clinical Groups | Dermatology Group |
| Ying Wu | Pfizer, New York, NY, United States | Clinical Groups | Dermatology Group |
| Pirkko Pussinen | Hospital District of Helsinki and Uusimaa, Helsinki, Finland | Clinical Groups | Odontology Group |
| Aino Salminen | Hospital District of Helsinki and Uusimaa, Helsinki, Finland | Clinical Groups | Odontology Group |
| Tuula Salo | Hospital District of Helsinki and Uusimaa, Helsinki, Finland | Clinical Groups | Odontology Group |
| David Rice | Hospital District of Helsinki and Uusimaa, Helsinki, Finland | Clinical Groups | Odontology Group |
| Pekka Nieminen | Hospital District of Helsinki and Uusimaa, Helsinki, Finland | Clinical Groups | Odontology Group |
| Ulla Palotie | Hospital District of Helsinki and Uusimaa, Helsinki, Finland | Clinical Groups | Odontology Group |
| Maria Siponen | Northern Savo Hospital District, Kuopio, Finland | Clinical Groups | Odontology Group |
| Liisa Suominen | Northern Savo Hospital District, Kuopio, Finland | Clinical Groups | Odontology Group |
| Päivi Mäntylä | Northern Savo Hospital District, Kuopio, Finland | Clinical Groups | Odontology Group |
| Ulvi Gursoy | Hospital District of Southwest Finland, Turku, Finland | Clinical Groups | Odontology Group |
| Vuokko Anttonen | Northern Ostrobothnia Hospital District, Oulu, Finland | Clinical Groups | Odontology Group |
| Kirsi Sipilä | Research Unit of Oral Health Sciences Faculty of Medicine, University of Oulu, Oulu, Finland; Medical Research Center, Oulu, Oulu University Hospital and University of Oulu, Oulu, Finland | Clinical Groups | Odontology Group |
| Rion Pendergrass | Genentech, San Francisco, CA, United States | Clinical Groups | Odontology Group |
| Hannele Laivuori | Institute for Molecular Medicine Finland (FIMM), HiLIFE, University of Helsinki, Helsinki, Finland | Clinical Groups | Women’s Health and Reproduction Group |
| Venla Kurra | Pirkanmaa Hospital District, Tampere, Finland | Clinical Groups | Women’s Health and Reproduction Group |
| Laura Kotaniemi-Talonen | Pirkanmaa Hospital District, Tampere, Finland | Clinical Groups | Women’s Health and Reproduction Group |
| Oskari Heikinheimo | Hospital District of Helsinki and Uusimaa, Helsinki, Finland | Clinical Groups | Women’s Health and Reproduction Group |
| Ilkka Kalliala | Hospital District of Helsinki and Uusimaa, Helsinki, Finland | Clinical Groups | Women’s Health and Reproduction Group |
| Lauri Aaltonen | Hospital District of Helsinki and Uusimaa, Helsinki, Finland | Clinical Groups | Women’s Health and Reproduction Group |
| Varpu Jokimaa | Hospital District of Southwest Finland, Turku, Finland | Clinical Groups | Women’s Health and Reproduction Group |
| Johannes Kettunen | Northern Ostrobothnia Hospital District, Oulu, Finland | Clinical Groups | Women’s Health and Reproduction Group |
| Marja Vääräsmäki | Northern Ostrobothnia Hospital District, Oulu, Finland | Clinical Groups | Women’s Health and Reproduction Group |
| Outi Uimari | Northern Ostrobothnia Hospital District, Oulu, Finland | Clinical Groups | Women’s Health and Reproduction Group |
| Laure Morin-Papunen | Northern Ostrobothnia Hospital District, Oulu, Finland | Clinical Groups | Women’s Health and Reproduction Group |
| Maarit Niinimäki | Northern Ostrobothnia Hospital District, Oulu, Finland | Clinical Groups | Women’s Health and Reproduction Group |
| Terhi Piltonen | Northern Ostrobothnia Hospital District, Oulu, Finland | Clinical Groups | Women’s Health and Reproduction Group |
| Katja Kivinen | Institute for Molecular Medicine Finland (FIMM), HiLIFE, University of Helsinki, Helsinki, Finland | Clinical Groups | Women’s Health and Reproduction Group |
| Elisabeth Widen | Institute for Molecular Medicine Finland (FIMM), HiLIFE, University of Helsinki, Helsinki, Finland | Clinical Groups | Women’s Health and Reproduction Group |
| Taru Tukiainen | Institute for Molecular Medicine Finland (FIMM), HiLIFE, University of Helsinki, Helsinki, Finland | Clinical Groups | Women’s Health and Reproduction Group |
| Mary Pat Reeve | Institute for Molecular Medicine Finland (FIMM), HiLIFE, University of Helsinki, Helsinki, Finland | Clinical Groups | Women’s Health and Reproduction Group |
| Mark Daly | Institute for Molecular Medicine Finland (FIMM), HiLIFE, University of Helsinki, Helsinki, Finland; Broad Institute of MIT and Harvard; Massachusetts General Hospital | Clinical Groups | Women’s Health and Reproduction Group |
| Niko Välimäki | University of Helsinki, Helsinki, Finland | Clinical Groups | Women’s Health and Reproduction Group |
| Eija Laakkonen | University of Jyväskylä, Jyväskylä, Finland | Clinical Groups | Women’s Health and Reproduction Group |
| Jaakko Tyrmi | University of Oulu, Oulu, Finland / University of Tampere, Tampere, Finland | Clinical Groups | Women’s Health and Reproduction Group |
| Heidi Silven | University of Oulu, Oulu, Finland | Clinical Groups | Women’s Health and Reproduction Group |
| Eeva Sliz | University of Oulu, Oulu, Finland | Clinical Groups | Women’s Health and Reproduction Group |
| Riikka Arffman | University of Oulu, Oulu, Finland | Clinical Groups | Women’s Health and Reproduction Group |
| Susanna Savukoski | University of Oulu, Oulu, Finland | Clinical Groups | Women’s Health and Reproduction Group |
| Triin Laisk | Estonian biobank, Tartu, Estonia | Clinical Groups | Women’s Health and Reproduction Group |
| Natalia Pujol | Estonian biobank, Tartu, Estonia | Clinical Groups | Women’s Health and Reproduction Group |
| Mengzhen Liu | Abbvie, Chicago, IL, United States | Clinical Groups | Women’s Health and Reproduction Group |
| Bridget Riley-Gillis | Abbvie, Chicago, IL, United States | Clinical Groups | Women’s Health and Reproduction Group |
| Rion Pendergrass | Genentech, San Francisco, CA, United States | Clinical Groups | Women’s Health and Reproduction Group |
| Janet Kumar | GlaxoSmithKline, Collegeville, PA, United States | Clinical Groups | Women’s Health and Reproduction Group |
| Kirsi Auro | GlaxoSmithKline, Espoo, Finland | Clinical Groups | Women’s Health and Reproduction Group |
| Iiris Hovatta | University of Helsinki, Finland | Clinical Groups | Depression group |
| Chia-Yen Chen | Biogen, Cambridge, MA, United States | Clinical Groups | Depression group |
| Erkki Isometsä | Hospital District of Helsinki and Uusimaa, Helsinki, Finland | Clinical Groups | Depression group |
| Hanna Ollila | Institute for Molecular Medicine Finland (FIMM), HiLIFE, University of Helsinki, Helsinki, Finland | Clinical Groups | Depression group |
| Jaana Suvisaari | Finnish Institute for Health and Welfare (THL), Helsinki, Finland | Clinical Groups | Depression group |
| Antti Mäkitie | Department of Otorhinolaryngology - Head and Neck Surgery, University of Helsinki and Helsinki University Hospital, Helsinki, Finland | Clinical Groups | ENT (ear, nose and throath) Group |
| Argyro Bizaki-Vallaskangas | Pirkanmaa Hospital District, Tampere, Finland | Clinical Groups | ENT (ear, nose and throath) Group |
| Sanna Toppila-Salmi | University of Eastern Finland and Kuopio University Hospital, Department of Otorhinolaryngology, Kuopio, Finland and Department of Allergy, Helsinki University Hospital and University of Helsinki, Finland | Clinical Groups | ENT (ear, nose and throath) Group |
| Tytti Willberg | Hospital District of Southwest Finland, Turku, Finland | Clinical Groups | ENT (ear, nose and throath) Group |
| Elmo Saarentaus | Institute for Molecular Medicine Finland (FIMM), HiLIFE, University of Helsinki, Helsinki, Finland | Clinical Groups | ENT (ear, nose and throath) Group |
| Antti Aarnisalo | Hospital District of Helsinki and Uusimaa, Helsinki, Finland | Clinical Groups | ENT (ear, nose and throath) Group |
| Eveliina Salminen | Hospital District of Helsinki and Uusimaa, Helsinki, Finland | Clinical Groups | ENT (ear, nose and throath) Group |
| Elisa Rahikkala | Northern Ostrobothnia Hospital District, Oulu, Finland | Clinical Groups | ENT (ear, nose and throath) Group |
| Johannes Kettunen | Northern Ostrobothnia Hospital District, Oulu, Finland | Clinical Groups | ENT (ear, nose and throath) Group |
| Kristiina Aittomäki | Department of Medical Genetics, Helsinki University Central Hospital, Helsinki, Finland | Clinical Groups | POI (premature ovarian failure) Group |
| Fredrik Åberg | Transplantation and Liver Surgery Clinic, Helsinki University Hospital, Helsinki University, Helsinki, Finland | Clinical Groups | LiverScore Group |
| Mitja Kurki | Institute for Molecular Medicine Finland (FIMM), HiLIFE, University of Helsinki, Helsinki, Finland; Broad Institute, Cambridge, MA, United States | FinnGen Analysis working group | FinnGen Analysis working group |
| Samuli Ripatti | Institute for Molecular Medicine Finland (FIMM), HiLIFE, University of Helsinki, Helsinki, Finland | FinnGen Analysis working group | FinnGen Analysis working group |
| Mark Daly | Institute for Molecular Medicine, Finland (FIMM), HiLIFE, University of Helsinki, Helsinki, Finland; Broad Institute of MIT and Harvard; Massachusetts General Hospital | FinnGen Analysis working group | FinnGen Analysis working group |
| Juha Karjalainen | Institute for Molecular Medicine Finland (FIMM), HiLIFE, University of Helsinki, Helsinki, Finland | FinnGen Analysis working group | FinnGen Analysis working group |
| Aki Havulinna | Institute for Molecular Medicine Finland (FIMM), HiLIFE, University of Helsinki, Helsinki, Finland; Finnish Institute for Health and Welfare (THL), Helsinki, Finland | FinnGen Analysis working group | FinnGen Analysis working group |
| Juha Mehtonen | Institute for Molecular Medicine Finland (FIMM), HiLIFE, University of Helsinki, Helsinki, Finland | FinnGen Analysis working group | FinnGen Analysis working group |
| Priit Palta | Institute for Molecular Medicine Finland (FIMM), HiLIFE, University of Helsinki, Helsinki, Finland | FinnGen Analysis working group | FinnGen Analysis working group |
| Shabbeer Hassan | Institute for Molecular Medicine Finland (FIMM), HiLIFE, University of Helsinki, Helsinki, Finland | FinnGen Analysis working group | FinnGen Analysis working group |
| Pietro Della Briotta Parolo | Institute for Molecular Medicine Finland (FIMM), HiLIFE, University of Helsinki, Helsinki, Finland | FinnGen Analysis working group | FinnGen Analysis working group |
| Wei Zhou | Broad Institute, Cambridge, MA, United States | FinnGen Analysis working group | FinnGen Analysis working group |
| Mutaamba Maasha | Broad Institute, Cambridge, MA, United States | FinnGen Analysis working group | FinnGen Analysis working group |
| Shabbeer Hassan | Institute for Molecular Medicine Finland (FIMM), HiLIFE, University of Helsinki, Helsinki, Finland | FinnGen Analysis working group | FinnGen Analysis working group |
| Susanna Lemmelä | Institute for Molecular Medicine Finland (FIMM), HiLIFE, University of Helsinki, Helsinki, Finland | FinnGen Analysis working group | FinnGen Analysis working group |
| Manuel Rivas | University of Stanford, Stanford, CA, United States | FinnGen Analysis working group | FinnGen Analysis working group |
| Aarno Palotie | Institute for Molecular Medicine Finland (FIMM), HiLIFE, University of Helsinki, Helsinki, Finland | FinnGen Analysis working group | FinnGen Analysis working group |
| Aoxing Liu | Institute for Molecular Medicine Finland (FIMM), HiLIFE, University of Helsinki, Helsinki, Finland | FinnGen Analysis working group | FinnGen Analysis working group |
| Arto Lehisto | Institute for Molecular Medicine Finland (FIMM), HiLIFE, University of Helsinki, Helsinki, Finland | FinnGen Analysis working group | FinnGen Analysis working group |
| Andrea Ganna | Institute for Molecular Medicine Finland (FIMM), HiLIFE, University of Helsinki, Helsinki, Finland | FinnGen Analysis working group | FinnGen Analysis working group |
| Vincent Llorens | Institute for Molecular Medicine Finland (FIMM), HiLIFE, University of Helsinki, Helsinki, Finland | FinnGen Analysis working group | FinnGen Analysis working group |
| Hannele Laivuori | Institute for Molecular Medicine Finland (FIMM), HiLIFE, University of Helsinki, Helsinki, Finland | FinnGen Analysis working group | FinnGen Analysis working group |
| Taru Tukiainen | Institute for Molecular Medicine Finland (FIMM), HiLIFE, University of Helsinki, Helsinki, Finland | FinnGen Analysis working group | FinnGen Analysis working group |
| Mary Pat Reeve | Institute for Molecular Medicine Finland (FIMM), HiLIFE, University of Helsinki, Helsinki, Finland | FinnGen Analysis working group | FinnGen Analysis working group |
| Henrike Heyne | Institute for Molecular Medicine Finland (FIMM), HiLIFE, University of Helsinki, Helsinki, Finland | FinnGen Analysis working group | FinnGen Analysis working group |
| Nina Mars | Institute for Molecular Medicine Finland (FIMM), HiLIFE, University of Helsinki, Helsinki, Finland | FinnGen Analysis working group | FinnGen Analysis working group |
| Joel Rämö | Institute for Molecular Medicine Finland (FIMM), HiLIFE, University of Helsinki, Helsinki, Finland | FinnGen Analysis working group | FinnGen Analysis working group |
| Elmo Saarentaus | Institute for Molecular Medicine Finland (FIMM), HiLIFE, University of Helsinki, Helsinki, Finland | FinnGen Analysis working group | FinnGen Analysis working group |
| Hanna Ollila | Institute for Molecular Medicine Finland (FIMM), HiLIFE, University of Helsinki, Helsinki, Finland | FinnGen Analysis working group | FinnGen Analysis working group |
| Rodos Rodosthenous | Institute for Molecular Medicine Finland (FIMM), HiLIFE, University of Helsinki, Helsinki, Finland | FinnGen Analysis working group | FinnGen Analysis working group |
| Satu Strausz | Institute for Molecular Medicine Finland (FIMM), HiLIFE, University of Helsinki, Helsinki, Finland | FinnGen Analysis working group | FinnGen Analysis working group |
| Tuula Palotie | University of Helsinki and Hospital District of Helsinki and Uusimaa, Helsinki, Finland | FinnGen Analysis working group | FinnGen Analysis working group |
| Kimmo Palin | University of Helsinki, Helsinki, Finland | FinnGen Analysis working group | FinnGen Analysis working group |
| Javier Garcia-Tabuenca | University of Tampere, Tampere, Finland | FinnGen Analysis working group | FinnGen Analysis working group |
| Harri Siirtola | University of Tampere, Tampere, Finland | FinnGen Analysis working group | FinnGen Analysis working group |
| Tuomo Kiiskinen | Institute for Molecular Medicine Finland (FIMM), HiLIFE, University of Helsinki, Helsinki, Finland | FinnGen Analysis working group | FinnGen Analysis working group |
| Jiwoo Lee | Institute for Molecular Medicine Finland (FIMM), HiLIFE, University of Helsinki, Helsinki, Finland; Broad Institute, Cambridge, MA, United States | FinnGen Analysis working group | FinnGen Analysis working group |
| Kristin Tsuo | Institute for Molecular Medicine Finland (FIMM), HiLIFE, University of Helsinki, Helsinki, Finland; Broad Institute, Cambridge, MA, United States | FinnGen Analysis working group | FinnGen Analysis working group |
| Amanda Elliott | Institute for Molecular Medicine Finland (FIMM), HiLIFE, University of Helsinki, Helsinki, Finland; Broad Institute, Cambridge, MA, USA and Massachusetts General Hospital, Boston, MA, USA | FinnGen Analysis working group | FinnGen Analysis working group |
| Kati Kristiansson | THL Biobank / Finnish Institute for Health and Welfare (THL), Helsinki, Finland | FinnGen Analysis working group | FinnGen Analysis working group |
| Mikko Arvas | Finnish Red Cross Blood Service / Finnish Hematology Registry and Clinical Biobank, Helsinki, Finland | FinnGen Analysis working group | FinnGen Analysis working group |
| Kati Hyvärinen | Finnish Red Cross Blood Service, Helsinki, Finland | FinnGen Analysis working group | FinnGen Analysis working group |
| Jarmo Ritari | Finnish Red Cross Blood Service, Helsinki, Finland | FinnGen Analysis working group | FinnGen Analysis working group |
| Olli Carpén | Helsinki Biobank / Helsinki University and Hospital District of Helsinki and Uusimaa, Helsinki | FinnGen Analysis working group | FinnGen Analysis working group |
| Johannes Kettunen | Northern Finland Biobank Borealis / University of Oulu / Northern Ostrobothnia Hospital District, Oulu, Finland | FinnGen Analysis working group | FinnGen Analysis working group |
| Katri Pylkäs | University of Oulu, Oulu, Finland | FinnGen Analysis working group | FinnGen Analysis working group |
| Eeva Sliz | University of Oulu, Oulu, Finland | FinnGen Analysis working group | FinnGen Analysis working group |
| Minna Karjalainen | University of Oulu, Oulu, Finland | FinnGen Analysis working group | FinnGen Analysis working group |
| Tuomo Mantere | Northern Finland Biobank Borealis / University of Oulu / Northern Ostrobothnia Hospital District, Oulu, Finland | FinnGen Analysis working group | FinnGen Analysis working group |
| Eeva Kangasniemi | Finnish Clinical Biobank Tampere / University of Tampere / Pirkanmaa Hospital District, Tampere, Finland | FinnGen Analysis working group | FinnGen Analysis working group |
| Sami Heikkinen | University of Eastern Finland, Kuopio, Finland | FinnGen Analysis working group | FinnGen Analysis working group |
| Arto Mannermaa | Biobank of Eastern Finland / University of Eastern Finland / Northern Savo Hospital District, Kuopio, Finland | FinnGen Analysis working group | FinnGen Analysis working group |
| Eija Laakkonen | University of Jyväskylä, Jyväskylä, Finland | FinnGen Analysis working group | FinnGen Analysis working group |
| Nina Pitkänen | Auria Biobank / University of Turku / Hospital District of Southwest Finland, Turku, Finland | FinnGen Analysis working group | FinnGen Analysis working group |
| Samuel Lessard | Translational Sciences, Sanofi R&D, Framingham, MA, USA | FinnGen Analysis working group | FinnGen Analysis working group |
| Clément Chatelain | Translational Sciences, Sanofi R&D, Framingham, MA, USA | FinnGen Analysis working group | FinnGen Analysis working group |
| Lila Kallio | Auria Biobank / University of Turku / Hospital District of Southwest Finland, Turku, Finland | Biobank directors | Biobank directors |
| Tiina Wahlfors | THL Biobank / Finnish Institute for Health and Welfare (THL), Helsinki, Finland | Biobank directors | Biobank directors |
| Jukka Partanen | Finnish Red Cross Blood Service / Finnish Hematology Registry and Clinical Biobank, Helsinki, Finland | Biobank directors | Biobank directors |
| Eero Punkka | Helsinki Biobank / Helsinki University and Hospital District of Helsinki and Uusimaa, Helsinki | Biobank directors | Biobank directors |
| Raisa Serpi | Northern Finland Biobank Borealis / University of Oulu / Northern Ostrobothnia Hospital District, Oulu, Finland | Biobank directors | Biobank directors |
| Sanna Siltanen | Finnish Clinical Biobank Tampere / University of Tampere / Pirkanmaa Hospital District, Tampere, Finland | Biobank directors | Biobank directors |
| Veli-Matti Kosma | Biobank of Eastern Finland / University of Eastern Finland / Northern Savo Hospital District, Kuopio, Finland | Biobank directors | Biobank directors |
| Teijo Kuopio | Central Finland Biobank / University of Jyväskylä / Central Finland Health Care District, Jyväskylä, Finland | Biobank directors | Biobank directors |
| Anu Jalanko | Institute for Molecular Medicine Finland (FIMM), HiLIFE, University of Helsinki, Helsinki, Finland | FinnGen Teams | Administration |
| Huei-Yi Shen | Institute for Molecular Medicine Finland (FIMM), HiLIFE, University of Helsinki, Helsinki, Finland | FinnGen Teams | Administration |
| Risto Kajanne | Institute for Molecular Medicine Finland (FIMM), HiLIFE, University of Helsinki, Helsinki, Finland | FinnGen Teams | Administration |
| Mervi Aavikko | Institute for Molecular Medicine Finland (FIMM), HiLIFE, University of Helsinki, Helsinki, Finland | FinnGen Teams | Administration |
| Helen Cooper | Institute for Molecular Medicine Finland (FIMM), HiLIFE, University of Helsinki, Helsinki, Finland | FinnGen Teams | Administration |
| Denise Öller | Institute for Molecular Medicine Finland (FIMM), HiLIFE, University of Helsinki, Helsinki, Finland | FinnGen Teams | Administration |
| Rasko Leinonen | Institute for Molecular Medicine Finland (FIMM), HiLIFE, University of Helsinki, Helsinki, Finland; European Molecular Biology Laboratory, European Bioinformatics Institute, Cambridge, UK | FinnGen Teams | Administration |
| Henna Palin | Finnish Clinical Biobank Tampere / University of Tampere / Pirkanmaa Hospital District, Tampere, Finland | FinnGen Teams | Administration |
| Malla-Maria Linna | Helsinki Biobank / Helsinki University and Hospital District of Helsinki and Uusimaa, Helsinki | FinnGen Teams | Administration |
| Mitja Kurki | Institute for Molecular Medicine Finland (FIMM), HiLIFE, University of Helsinki, Helsinki, Finland; Broad Institute, Cambridge, MA, United States | FinnGen Teams | Analysis |
| Juha Karjalainen | Institute for Molecular Medicine Finland (FIMM), HiLIFE, University of Helsinki, Helsinki, Finland | FinnGen Teams | Analysis |
| Pietro Della Briotta Parolo | Institute for Molecular Medicine Finland (FIMM), HiLIFE, University of Helsinki, Helsinki, Finland | FinnGen Teams | Analysis |
| Arto Lehisto | Institute for Molecular Medicine Finland (FIMM), HiLIFE, University of Helsinki, Helsinki, Finland | FinnGen Teams | Analysis |
| Juha Mehtonen | Institute for Molecular Medicine Finland (FIMM), HiLIFE, University of Helsinki, Helsinki, Finland | FinnGen Teams | Analysis |
| Wei Zhou | Broad Institute, Cambridge, MA, United States | FinnGen Teams | Analysis |
| Masahiro Kanai | Broad Institute, Cambridge, MA, United States | FinnGen Teams | Analysis |
| Mutaamba Maasha | Broad Institute, Cambridge, MA, United States | FinnGen Teams | Analysis |
| Zhili Zheng | Broad Institute, Cambridge, MA, United States | FinnGen Teams | Analysis |
| Hannele Laivuori | Institute for Molecular Medicine Finland (FIMM), HiLIFE, University of Helsinki, Helsinki, Finland | FinnGen Teams | Clinical Endpoint Development |
| Aki Havulinna | Institute for Molecular Medicine Finland (FIMM), HiLIFE, University of Helsinki, Helsinki, Finland; Finnish Institute for Health and Welfare (THL), Helsinki, Finland | FinnGen Teams | Clinical Endpoint Development |
| Susanna Lemmelä | Institute for Molecular Medicine Finland (FIMM), HiLIFE, University of Helsinki, Helsinki, Finland | FinnGen Teams | Clinical Endpoint Development |
| Tuomo Kiiskinen | Institute for Molecular Medicine Finland (FIMM), HiLIFE, University of Helsinki, Helsinki, Finland | FinnGen Teams | Clinical Endpoint Development |
| L. Elisa Lahtela | Institute for Molecular Medicine Finland (FIMM), HiLIFE, University of Helsinki, Helsinki, Finland | FinnGen Teams | Clinical Endpoint Development |
| Mari Kaunisto | Institute for Molecular Medicine Finland (FIMM), HiLIFE, University of Helsinki, Helsinki, Finland | FinnGen Teams | Communication |
| Elina Kilpeläinen | Institute for Molecular Medicine Finland (FIMM), HiLIFE, University of Helsinki, Helsinki, Finland | FinnGen Teams | E-Science |
| Timo P. Sipilä | Institute for Molecular Medicine Finland (FIMM), HiLIFE, University of Helsinki, Helsinki, Finland | FinnGen Teams | E-Science |
| Oluwaseun Alexander Dada | Institute for Molecular Medicine Finland (FIMM), HiLIFE, University of Helsinki, Helsinki, Finland | FinnGen Teams | E-Science |
| Awaisa Ghazal | Institute for Molecular Medicine Finland (FIMM), HiLIFE, University of Helsinki, Helsinki, Finland | FinnGen Teams | E-Science |
| Anastasia Kytölä | Institute for Molecular Medicine Finland (FIMM), HiLIFE, University of Helsinki, Helsinki, Finland | FinnGen Teams | E-Science |
| Rigbe Weldatsadik | Institute for Molecular Medicine Finland (FIMM), HiLIFE, University of Helsinki, Helsinki, Finland | FinnGen Teams | E-Science |
| Sanni Ruotsalainen | Institute for Molecular Medicine Finland (FIMM), HiLIFE, University of Helsinki, Helsinki, Finland | FinnGen Teams | E-Science |
| Kati Donner | Institute for Molecular Medicine Finland (FIMM), HiLIFE, University of Helsinki, Helsinki, Finland | FinnGen Teams | Genotyping |
| Timo P. Sipilä | Institute for Molecular Medicine Finland (FIMM), HiLIFE, University of Helsinki, Helsinki, Finland | FinnGen Teams | Genotyping |
| Anu Loukola | Helsinki Biobank / Helsinki University and Hospital District of Helsinki and Uusimaa, Helsinki | FinnGen Teams | Sample Collection Coordination |
| Päivi Laiho | THL Biobank / Finnish Institute for Health and Welfare (THL), Helsinki, Finland | FinnGen Teams | Sample Logistics |
| Tuuli Sistonen | THL Biobank / Finnish Institute for Health and Welfare (THL), Helsinki, Finland | FinnGen Teams | Sample Logistics |
| Essi Kaiharju | THL Biobank / Finnish Institute for Health and Welfare (THL), Helsinki, Finland | FinnGen Teams | Sample Logistics |
| Markku Laukkanen | THL Biobank / Finnish Institute for Health and Welfare (THL), Helsinki, Finland | FinnGen Teams | Sample Logistics |
| Elina Järvensivu | THL Biobank / Finnish Institute for Health and Welfare (THL), Helsinki, Finland | FinnGen Teams | Sample Logistics |
| Sini Lähteenmäki | THL Biobank / Finnish Institute for Health and Welfare (THL), Helsinki, Finland | FinnGen Teams | Sample Logistics |
| Lotta Männikkö | THL Biobank / Finnish Institute for Health and Welfare (THL), Helsinki, Finland | FinnGen Teams | Sample Logistics |
| Regis Wong | THL Biobank / Finnish Institute for Health and Welfare (THL), Helsinki, Finland | FinnGen Teams | Sample Logistics |
| Auli Toivola | THL Biobank / Finnish Institute for Health and Welfare (THL), Helsinki, Finland | FinnGen Teams | Sample Logistics |
| Minna Brunfeldt | THL Biobank / Finnish Institute for Health and Welfare (THL), Helsinki, Finland | FinnGen Teams | Registry Data Operations |
| Hannele Mattsson | THL Biobank / Finnish Institute for Health and Welfare (THL), Helsinki, Finland | FinnGen Teams | Registry Data Operations |
| Kati Kristiansson | THL Biobank / Finnish Institute for Health and Welfare (THL), Helsinki, Finland | FinnGen Teams | Registry Data Operations |
| Susanna Lemmelä | Institute for Molecular Medicine Finland (FIMM), HiLIFE, University of Helsinki, Helsinki, Finland | FinnGen Teams | Registry Data Operations |
| Sami Koskelainen | THL Biobank / Finnish Institute for Health and Welfare (THL), Helsinki, Finland | FinnGen Teams | Registry Data Operations |
| Tero Hiekkalinna | THL Biobank / Finnish Institute for Health and Welfare (THL), Helsinki, Finland | FinnGen Teams | Registry Data Operations |
| Teemu Paajanen | THL Biobank / Finnish Institute for Health and Welfare (THL), Helsinki, Finland | FinnGen Teams | Registry Data Operations |
| Priit Palta | Institute for Molecular Medicine Finland (FIMM), HiLIFE, University of Helsinki, Helsinki, Finland | FinnGen Teams | Sequencing Informatics |
| Shuang Luo | Institute for Molecular Medicine Finland (FIMM), HiLIFE, University of Helsinki, Helsinki, Finland | FinnGen Teams | Sequencing Informatics |
| Tarja Laitinen | Pirkanmaa Hospital District, Tampere, Finland | FinnGen Teams | Trajectory |
| Mary Pat Reeve | Institute for Molecular Medicine Finland (FIMM), HiLIFE, University of Helsinki, Helsinki, Finland | FinnGen Teams | Trajectory |
| Shanmukha Sampath Padmanabhuni | Institute for Molecular Medicine Finland (FIMM), HiLIFE, University of Helsinki, Helsinki, Finland | FinnGen Teams | Trajectory |
| Marianna Niemi | University of Tampere, Tampere, Finland | FinnGen Teams | Trajectory |
| Harri Siirtola | University of Tampere, Tampere, Finland | FinnGen Teams | Trajectory |
| Javier Gracia-Tabuenca | University of Tampere, Tampere, Finland | FinnGen Teams | Trajectory |
| Mika Helminen | University of Tampere, Tampere, Finland | FinnGen Teams | Trajectory |
| Tiina Luukkaala | University of Tampere, Tampere, Finland | FinnGen Teams | Trajectory |
| Iida Vähätalo | University of Tampere, Tampere, Finland | FinnGen Teams | Trajectory |
| Jyrki Tammerluoto | Institute for Molecular Medicine Finland (FIMM), HiLIFE, University of Helsinki, Helsinki, Finland | FinnGen Teams | Data protection officer |
| Marco Hautalahti | Finnish Biobank Cooperative - FINBB | FinnGen Teams | FINBB - Finnish biobank cooperative |
| Johanna Mäkelä | Finnish Biobank Cooperative - FINBB | FinnGen Teams | FINBB - Finnish biobank cooperative |
| Sarah Smith | Finnish Biobank Cooperative - FINBB | FinnGen Teams | FINBB - Finnish biobank cooperative |
| Tom Southerington | Finnish Biobank Cooperative - FINBB | FinnGen Teams | FINBB - Finnish biobank cooperative |
| Petri Lehto | Finnish Biobank Cooperative - FINBB | FinnGen Teams | FINBB - Finnish biobank cooperative |
